# An Allele of the MTHFR one-carbon metabolism gene predicts severity of COVID-19

**DOI:** 10.1101/2025.02.28.25323089

**Authors:** Boryana Petrova, Caitlin Syphurs, Andrew J Culhane, Jing Chen, Ernie Chen, Chris Cotsapas, Denise Esserman, Ruth Montgomery, Steven Kleinstein, Kinga Smolen, Kevin Mendez, IMPACC Network, Jessica Lasky-Su, Hanno Steen, Ofer Levy, Joann Diray-Arce, Naama Kanarek

**Author notes:** (equal contribution) (Boryana Petrova, Joann Diray-Arce, Naama Kanarek).

## Abstract

While the public health burden of SARS-CoV-2 infection has lessened due to natural and vaccine-acquired immunity, the emergence of less virulent variants, and antiviral medications, COVID-19 continues to take a significant toll. There are > 10,000 new hospitalizations per week in the U.S., many of whom develop post-acute sequelae of SARS-CoV-2 (PASC), or “long COVID”, with long-term health issues and compromised quality of life. Early identification of individuals at high risk of severe COVID-19 is key for monitoring and supporting respiratory status and improving outcomes. Therefore, precision tools for early detection of patients at high risk of severe disease can reduce morbidity and mortality. Here we report an untargeted and longitudinal metabolomic study of plasma derived from adult patients with COVID-19. One-carbon metabolism, a pathway previously shown as critical for viral propagation and disease progression, and a potential target for COVID-19 treatment, scored strongly as differentially abundant in patients with severe COVID-19. A follow-up targeted metabolite profiling revealed that one arm of the one-carbon metabolism pathway, the methionine cycle, is a major driver of the metabolic profile associated with disease severity. The methionine cycle produces S-adenosylmethionine (SAM), the methyl group donor important for methylation of DNA, RNA, and proteins, and its high abundance was reported to correlate with disease severity. Further, genomic data from the profiled patients revealed a genetic contributor to methionine metabolism and identified the C677T allele of the *MTHFR* gene as a pre-existing predictor of disease trajectory - patients homozygous for the *MTHFR* C677T have higher incidence of experiencing severe disease. Our results raise the possibility that screening for the common genetic *MTHFR* variant may be an actionable approach to stratify risk of COVID severity and may inform novel precision COVID-19 treatment strategies.

## Introduction

While SARS-CoV-2 has lessened with the advent of vaccines, antivirals, and less pathogenic variants, COVID-19 continues to take a significant toll across the globe^1, 2^. There remains an unmet need to better predict who is at greatest risk of severe or long COVID-19 and to define relevant molecular pathways that may be amenable to novel drugs to reduce the impact of COVID-19.

Post-acute sequelae of SARS-CoV-2 (PASC), commonly referred to as long COVID-19, has emerged as a significant public health issue. PASC is particularly prevalent among hospitalized COVID-19 patients, with some studies reporting that up to 50% experience persistent physical, cognitive, or mental impairments months after their initial diagnosis^3^. The condition poses considerable challenges to quality of life, healthcare systems, and economic productivity^4^. Multiple risk factors and immune profiles during the acute phase of infection correlate with delayed clinical and functional recovery after hospital discharge^5, 6^.

Among the pathways that are relevant to host responses to infection is one-carbon metabolism (1CM)^7^ that encompasses metabolic pathways that involve the transfer of a one-carbon moiety from a donor molecule to an acceptor molecule.1CM enables catabolism and synthesis of key metabolites and cellular building blocks including a few amino acids (serine, glycine, glutamine, and histidine), and nucleotides^8^, 1CM is critical in supporting rapid cell proliferation and is a target of anti-cancer therapy^9^. Similar to rapidly proliferating cancer cells, virus-infected cells also shift their metabolism to support intense demand for nucleic acids to support viral RNA production^10^. Further, it was shown that COVID-19 infected cells rely on the host cell’s 1CM and are sensitive to its inhibition by the anti-folate methotrexate^7, 11^.

Methylenetetrahydrofolate reductase encoded by the gene *MTHFR* is a key enzyme responsible for the conversion of 5,10-methylene THF to 5-methyl THF. 5-methyl THF is a critical form of folate that feeds the methylation cycle for synthesis of methionine and the methyl donor S-adenosyl methionine (SAM). The *MTHFR* gene has been widely studied due to the common polymorphism C677T, found in ∼10% of individuals in North America, that causes conversion of valine to alanine, resulting in a hypomorph with reduced enzymatic activity^12^. Carriers of the hypomorph manifest increased plasma homocysteine levels, and association with several pathologies, including oncologic, vascular and metabolic diseases^13^.

The potential link between COVID-19 risk and *MTHFR* polymorphism was hypothesized early in the pandemic^14^. This hypothesis is further supported by a geographical correlation between COVID-19 spread and the prevalence of *MTHFR* polymorphism^15^, and the correlation between SAM levels and disease severity^16^. A few case reports also suggest associations between COVID-19 severity and specific complications such as neuroretinopathy or dermatological manifestations and *MTHFR* polymorphism^17–19^. Further, *MTHFR* polymorphisms and methylation status have also been investigated as long COVID-19 risk factors^20, 21^, though their utility in patient risk stratification remains uncertain.

We leveraged the NIH/NIAID-supported Immunophenotyping of a Coronavirus Cohort (IMPACC) cohort^22^ to investigate the relationship between COVID-19 progression and 1CM pathway intermediates. Targeted metabolomics revealed significant disruptions in the methionine cycle in severe COVID-19 cases. Integrating *MTHFR* C677T allele status and metabolic perturbations in 1CM pathway intermediates revealed potential predictors of severe COVID-19. Furthermore, we observed marked 1CM perturbations in PASC patients, and identified the combination of homozygous *MTHFR* C677T variant with a methionine-centered metabolite signature as a significant prediction tool for COVID-19 severity and PASC risk. This work supports future application of precision approaches employing integrated genomics and metabolomics to patient stratification for COVID-19 risk management.

## Results

### Metabolite profiling of early-collected COVID-19 plasma samples reveals correlation between one-carbon metabolism and disease severity

The IMPACC cohort represents a prospective longitudinal study that captures disease progression of 1,035 COVID-19 patients (Supplementary Table 1). Samples were collected across 2 phases: acute hospitalization phase and follow up post-hospitalization discharge. Each phase was subdivided into visits: within the hospitalization stage there were more frequent visits while there were longer time intervals between visits within the follow up phase (Figure 1A). A comprehensive range of clinical, biochemical, immunological and systems biology assays were performed during both segments as described previously^22^. Patients were retrospectively categorized into disease severity trajectories based on the severity of their clinical symptoms with Trajectory Group 5 being most severe and having a fatal outcome (Figure 1B).

**Figure 1.**
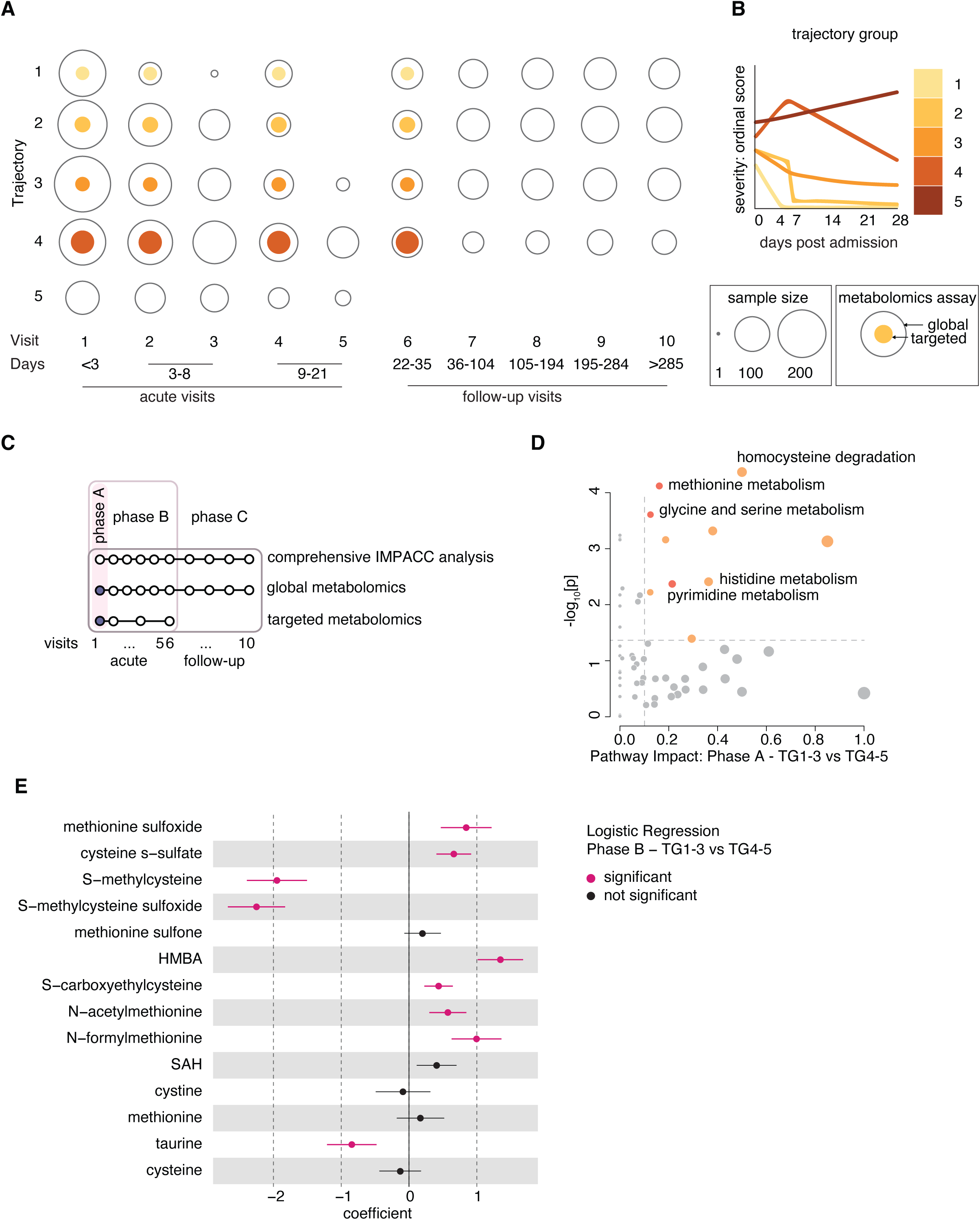
Integrated global and targeted plasma metabolomics of COVID-19 patients reveals one-carbon metabolism association with COVID-19 severity. A A schematic representing the IMPACC metabolomics cohorts according to trajectory groups and visit number. Days after admission are listed. Filled color-coded area within the circles represents the number of samples analyzed by targeted metabolomics, and open area outlines represent the number of samples analyzed by global metabolomics per visit and per disease trajectory group. B A schematic depicting the clinical trajectory group (TG) assignments (TG 1-5) for all IMPACC cohort participants (1,164 in total) as reported in Diray-Arce et al.^23^. The x-axis depicts days since hospital admission, while the y-axis displays the ordinal respiratory status score (lowest score indicates mild disease and discharge, highest score indicates mortality). C A schematic illustrating the study design and employed assays of the IMPACC cohort^22^ and outlining the scope of the global and targeted metabolomics assays and the separation into three distinct phases (A-C) encompassing sample collection during hospitalization (“acute”) and during follow-up (“follow up”). Highlighted is Phase A, that provided the data presented in panel B, and phase B, that is inclusive of phase A and that provided the data for panel E. D Pathway analysis of differential metabolites from targeted metabolomics profiling of phase A between mild (TG 1-3) and severe (TG 4-5) trajectories performed on the MetaboAnalyst platform, post log-transforming and Pareto scaling mean-centered data. Significantly different pathways (p < 0.05) are highlighted as follows: pathways with an impact between 0.1 and 1 are shown with orange circles, and those of particular interest are in dark orange. Significant pathways are marked with orange circles, while significant pathways of interest are dark orange. Other pathways are shown in grey. n=199 E Logistic regression analysis from global metabolomics of Phase B for methionine, cysteine, SAM and taurine metabolism pathway between mild (TG 1-3) and severe (TG 4-5) trajectory groups. Significantly different metabolites (p < 0.01, coefficient > 0) are highlighted in magenta. n=1055

Two different types of metabolomics assays were performed: global and targeted (Figure 1A, C and Figure S1A). Global metabolomics analysis captured all patients at all time points, while targeted metabolomics included a pre-selected subgroup. This subgroup included patients from three phases: phase A analyzed patients’ samples at Visit 1, phase B analyzed samples from Visit 1 through Visit 6 and phase C analyzed all samples within the IMPACC cohort including follow up samples (Figure 1C and S1A). Both global and targeted metabolomics analyses were used for hypothesis generation and validation. Global metabolomics assays were performed using the platform at Metabolon^23^ (Durham, NC), targeted metabolomic assays were performed at Boston Children’s Hospital (Boston, MA). Additional datasets such as clinical assessments or genomics, were accessible through the IMPACC network^5, 22, 24^.

To allow for the comparison and integration of data from two different metabolomics platforms, our study relied on metabolomics measurements which partially overlapped between the two platforms, allowing cross reference measurements per patient and visit and determination of a coefficient of correlation (Figure S1B). We compared three abundant metabolites, two of which were common blood indicators (creatine and creatinine) while the third had strong relevance to our study (methionine). All three metabolites showed strong correlation even though liquid chromatography mass spectrometry (LC-MS) methodology was not synchronized between the two platforms as this was precluded by use of proprietary methods for the global analysis by Metabolon. We performed batch normalization following previously published guidelines from Metabolon^25^, applying mean centering across the targeted metabolomics cohort, that successfully eliminated batch effects (Figure S1C).

As samples from Visit 1 became available, and in parallel to continued longitudinal sample collection in following visits, we defined phase A of our study (Figure 1C) and performed targeted metabolomics assay on 199 patient plasma samples collected at Visit 1. We performed pathway analysis of differential metabolites between disease severity trajectories: mild (Trajectory Groups 1-3) and severe (Trajectory Groups 4 and 5) (Figure 1D). This analysis highlighted several pathways involved in 1CM and utilization (methionine, histidine, glycine and serine, and homocysteine degradation), that were previously reported to correlate with various aspects of COVID-19 pathology viral replication^7, 10, 11, 26–28^. We next analyzed our global metabolomics dataset for data collected at phase B (Figure 1C). We performed logistic regression analysis that demonstrated significantly altered plasma metabolites in the methionine, cysteine, SAM, and taurine metabolism pathways between mild and severe disease trajectory groups (Figure 1E), and across all trajectory groups (Figure S1D). Therefore, our data from two independent metabolomics studies indicate early association between 1CM and COVID-19 disease severity and outcome, that remained significant at later time points. This implies that metabolic changes observed as early as Visit 1 are correlated with disease severity and outcome, suggesting potential early markers of disease trajectory or even a predisposition for disease severity rooted in the patient’s metabolic profile.

### Targeted metabolomics reveal methionine cycle alterations with COVID-19 disease progression and severity

Next, we studied what reactions within 1CM-related pathways drive the differential 1CM metabolite concentrations and their directionality in severe vs. mild COVID-19 patients. We detected metabolites from both the folate and methionine cycles (Figure 2A); The folate cycle drives nucleotide synthesis with 10-formyl THF as a carbon donor for purine synthesis and 5,10-methylene THF as a carbon donor for thymidylate synthesis^8^. The methionine cycle fuels methylation reactions through synthesis of SAM, the methyl group donor of many methyl transferases responsible for methylation of DNA, RNA, and proteins^8, 29^. Both global and targeted metabolomics analyses identified alterations in methionine levels in COVID-19 patient plasma (Figure 2B). Specifically, significant changes were observed between visits, while levels did not significantly vary between disease severity trajectory groups. Methionine levels first increased at Visit 2 and then decreased at Visits 4 and 6. This suggested correlation between methionine levels and COVID-19 disease progression. Further, these data shed light on the inconsistency in reporting methionine levels between studies that involve samples from patients with mild and severe disease, but that were collected at various times following disease onset^10^. However, measurements of methionine levels alone could not differentiate between disease severity trajectories at either visit. In contrast, the relative abundance of the methionine oxidation product methionine-sulfoxide was different between plasma samples from patients in different severity trajectory groups as well as throughout consecutive hospital visits especially in the more severe trajectories (TG4 and 5) (Figure 2C). These results were consistent in our targeted metabolomics cohort, despite the lower number of samples included in this subset, indicating that these results are platform independent.

**Figure 2.**
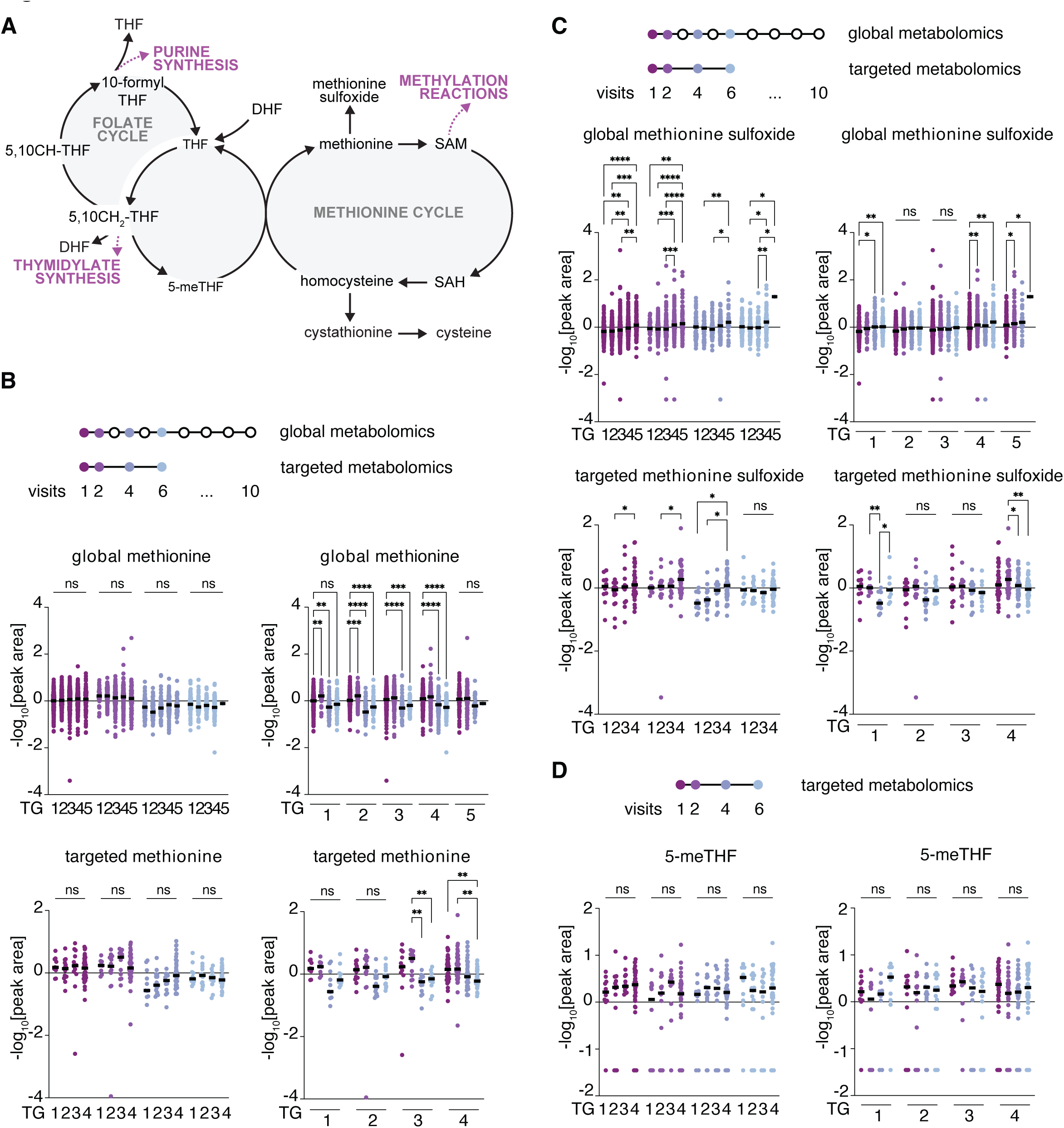
Relative changes in methionine metabolism are correlated with COVID-19 severity. A Schematic of one-carbon metabolism. Abbreviations are: DHF - dihydrofolate; THF - tetrahydrofolate; 5-meTHF – 5-methyl THF; 5,10-CH THF – 5,10-methenyl THF; 5,10-CH_2_ THF – 5,10-methylene THF; SAM – S-Adenosyl methionine; SAH – S-adenosylhomocysteine. Pathways downstream to the shown reactions are highlighted in magenta. B Methionine levels from global and targeted metabolomics. At the top a schematic to clarify samples included in the analysis and the visits color code. Below are metabolite profiling data that were mean-centered, log-transformed, and Pareto-scaled. Left or right panels are depicting the same data but organized either by trajectory groups (TG1-5) or by visit (V1,2,4 and 6) respectively. A two-way Anova (trajectory comparisons) or a mixed-effects (visit comparisons) analyses were performed, and corresponding p-values are indicated where *= p<0.01; **= p<0.001; ***= p<0.001; all remaining comparisons were not significant (ns) and were omitted for clarity. Global metabolomics n=2146; Targeted metabolomics n=199. TG – trajectory group. C As for B but depicting methionine sulfoxide levels. D 5-methyl THF (5-meTHF) levels from targeted metabolomics. At the top a schematic to clarify samples included in the analysis and the visits color code. Below are metabolite profiling data that were treated and represented as in B and C except 5-methyl THF was detected only in the targeted analysis. n=199

Beyond methionine and its oxidation product, we also analyzed the relative levels of other metabolites of the methionine cycle including glycine, serine, and SAH (Figure 2A). We noted significant differential levels in glycine between severity trajectory groups especially at Visits 2 and 4 (Figure S2A). Serine demonstrated more consistently significant differences between severity trajectory groups in all visits, with 1.5 – 1.85 fold decreased levels of serine with higher severity . Its relative levels were lower in more severe patients with for example a 1.8-fold decrease observed in TG5 vs TG1 at Visit 1 (Figure S2B). For SAH, significant differential levels were especially pronounced between trajectory group comparisons, while fewer changes were observed within trajectory per visit (Figure S2C). For glycine and serine the targeted assay results broadly aligned with global metabolomics findings but were underpowered to capture the biological variability, and SAH was not well-detected by targeted metabolomics for these samples.

Next, we performed targeted folate measurements, and detected relative plasma levels of the folate form most abundant in plasma, 5-methyl tetrahydrofolate (5-me THF). Despite there being some subtle changes between visits or trajectories, 5-me THF levels were not significantly different (Figure 2D). This suggests that the folate cycle may be less disrupted compared to the methionine cycle, or that 5-me THF homeostasis is more tightly maintained. However, we cannot rule out the possibility that our targeted assay is underpowered, and this negative result is influenced by the smaller number of samples available for targeted analysis. Overall, our data suggests that alterations in the methionine pathway, already observed at Visit 1, could be used to inform COVID-19 disease trajectory and outcome.

### Genetic susceptibility and alterations in the methionine pathway correlate with disease severity

We next addressed the possibility of a predisposition that might contribute to the differential levels of methionine cycle metabolites as early as early visit 1 (Figure 1D). A possible predisposition is genetic variance, that in the case of methionine metabolism can be the common variant of the *MTHFR* gene that encodes the MTHFR enzyme. MTHFR converts 5,10-methylene THF to 5-methyl THF, linking the folate and the methionine cycles (Figure S3A). Common genetic variants exist in the *MTHFR* gene, specifically, C677T (rs1801133) and A1298C (rs1801131) (Figure S3B). The more common heterozygous genotype (C/T, here denoted as GA to reflect the amino acid change) is present in ∼30-40% of the population worldwide. The homozygous mutant genotype (T/T, here denoted as AA) is present in ∼10-15% of individuals globally, and results in a substitution of alanine with valine at position 222 in the MTHFR enzyme (Figure S3B). This change is reported to reduce the enzyme’s activity to ∼30% of its normal levels with a reported impact on carriers’ physiology^13^. The A1298C variant also results in reduced enzyme activity, but to a lesser extent (70%, Figure S3B). Because *MTHFR* polymorphism can result in changes in the methionine cycle flux and levels of related metabolites^30^, we set to assess whether this common polymorphism plays a role in the methionine metabolite-related predisposition to severe COVID we identified.

We first verified that the *MTHFR* C677T allele distribution of the IMPACC cohort reflects the distribution found in the general population within the USA^31, 32^ (Figure S3C). Indeed, the IMPACC cohort had a very similar distribution of the MTHFR variants with 13.3% frequency of the AA allele, compared to the previously reported 14.5% in the general US population. We also found comparable allele frequency between severity trajectory groups (Figure S3D). Of note, in the cohort used for the targeted metabolomics analysis we identified a bias towards the AA allele that appeared at 18.4% frequency in Trajectory Group 4 and not found at all in the mild trajectory groups (Figure S3E). This might stem from the overrepresentation of samples in Trajectory Group 4 in this cohort (Figure 1A).

Next, we explored whether the C677T polymorphism could influence methionine metabolite status as early as study Visit 1. Because individuals with a specific *MTHFR* allele could be assessed as they present to the hospital, we wondered if *MTHFR* allele information and levels in the methionine cycle metabolites could be used to predict COVID-19 disease outcome and potentially serve as early biomarkers. To this end, we harnessed the IMPACC-wide clinical, genomic, and global metabolomic data (Figure 3A).

**Figure 3.**
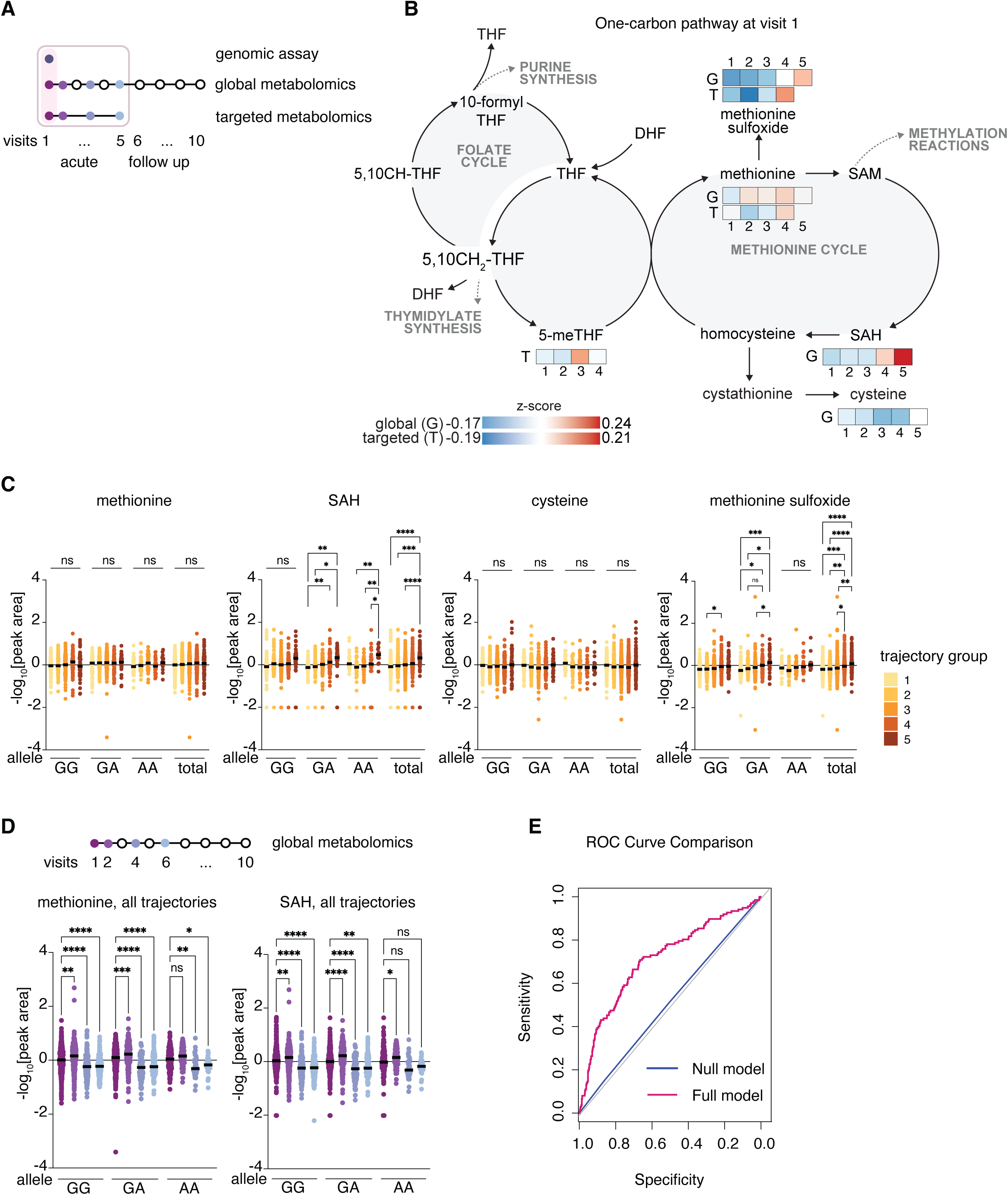
An increase in several metabolites in the methionine pathway at Visit 1 correlates with COVID-19 disease severity and modulation by *MTHFR* allele status. A A schematic depicting the visits and analyses of the IMPACC cohort used for data presented in the figure. Analyses include targeted metabolomics, global metabolomics, and genomic assay. B Schematic of one-carbon metabolism with overlaid differences in abundance between trajectory groups (TG, numbered 1-5) at visit 1 of individual metabolites detected either by global (G) or targeted (T) metabolite profiling. Differences in metabolite abundance between trajectory groups are represented as z-scores, indicated by a color code (see legend). Only metabolites that were detected by either global or targeted metabolomics analysis are represented. C Methionine, SAH, cysteine, and methionine sulfoxide levels detected by global metabolomics were stratified by *MTHFR* allele and disease trajectory group. Only data from visit 1 is depicted. Data were mean-centered, log-transformed, and Pareto-scaled. A mixed-effects analysis was performed, and corresponding p-values are indicated where *= p<0.01; **= p<0.001; ***= p<0.001; all remaining comparisons were not significant (ns) and were omitted for clarity. GG n = 510; GA n = 358; AA n = 133 D. Methionine and SAH levels detected by global metabolomics stratified by *MTHFR* allele and visit. Data were mean-centered, log-transformed, and Pareto-scaled. Data from all trajectories were combined. A mixed-effects analysis was performed, and corresponding p-values are indicated where *= p<0.01; **= p<0.001; ***= p<0.001; all remaining comparisons were not significant (ns). For methionine and SAH: V1, GG n = 480; V1, GA n = 345; V1, AA n = 129; V2, GG n = 304; V2, GA n = 204; V2, AA n = 82; V4, GG n = 164; V4, GA n = 116; V4, AA n = 36; V6, GG n = 153; V6, GA n = 102; V6, AA n = 31 E. ROC curve goodness of fit comparison Likelihood ratio test for null logistic model (predicting mortality based on *MTHFR* alleles) vs full logistic model (predicting mortality based on *MTHFR* alleles and baseline levels of the metabolites SAH, methionine and methionine-sulfoxide).

Among the methionine cycle metabolites that were detected in samples from Visit 1, methionine sulfoxide and SAH increased significantly with severity while methionine and cysteine were similar between trajectory groups (Figure 3B). When stratified by *MTHFR* allele, for methionine sulfoxide or SAH, there were differences between the individual disease susceptibilities. Namely, for SAH, patients with the AA allele seemed to show similar levels independent of disease trajectory. While for methionine-sulfoxide, significant differences were observed with the GA and AA allele and not the wildtype GG allele (Figure 3C). Changes in the abundance of these metabolites from Visit 1 to later visits were significant in all three genotypes, with lower number of participants likely driving loss of statistical power and reduced significance for the AA group (Figure 3D). Combination of the data for all trajectory groups together resulted in no significant difference in the levels of these metabolites when stratifying our results by *MTHFR* allele status per visit (Figure S3F). These results suggest that *MTHFR* allele status and methionine metabolism may serve as a predisposition and possibly informative biomarkers of COVID-19 disease severity. However, we were mostly underpowered to use our IMPACC cohort for analysis of *MTHFR* as a sole genetic driver of disease severity and mortality (Figure S4A), or a single parameter for significant hazard ratio, although we saw a trend (p = 0.077) towards a higher negative hazard ratio with one of the C677T hypomorphic alleles (Figure S4B).

Finally, we assessed if levels of individual metabolites together with the C677T *MTHFR* allele status at Visit 1 could serve as a prediction factor for mortality risk. We performed likelihood ratio analysis^33–35^. This is a statistical method that compares two competing logistic regression models: a null model (baseline, here - *MTHFR* C677T allele status only) and an alternative model (more complex, here - it includes baseline SAH, methionine and methionine sulfoxide levels in addition to *MTHFR* allele status), and tests whether the additional complexity of the alternative model significantly improves the fit to the data. The inclusion of the three metabolites in the model significantly improved the model’s ability to predict mortality compared to a model based solely on *MTHFR* allele genotype (Figure 3E). The Akaike information criterion (AIC) value decreased from 790.85 (null model) to 738.66 (full model) and the log-likelihood value increased from -392.43 to -363.33, indicating a better fit with the addition of metabolites while accounting for model complexity (□2(3)=58.2, □<0.001). In contrast, adding genetic information to a null model based only on baseline metabolites did not improve the model’s fit, likely because the genetic contribution is either fully captured by the metabolite data or provides limited additional predictive value in this context (Figure S4C). These findings suggest that the three metabolites provide additional predictive power beyond *MTHFR* status alone, highlighting their relevance in understanding disease severity.

### MTHFR allele status and early alterations in one-carbon metabolism provide prediction factor for long COVID-19

Next, we tested whether *MTHFR* allele status and early detectable aberrant 1CM can be used for predictions of post-acute sequelae of SARS-CoV-2 (PASC), that is a significant public health concern^36, 37^. For that, we used patient-reported outcomes (PROs) of IMPACC participants to stratify the IMPACC cohort by PASC severity, and we overlaid this data with *MTHFR* allele status and metabolite abundance of one-carbon metabolites detected by our global metabolomics analysis (Figure 4A).

**Figure 4.**
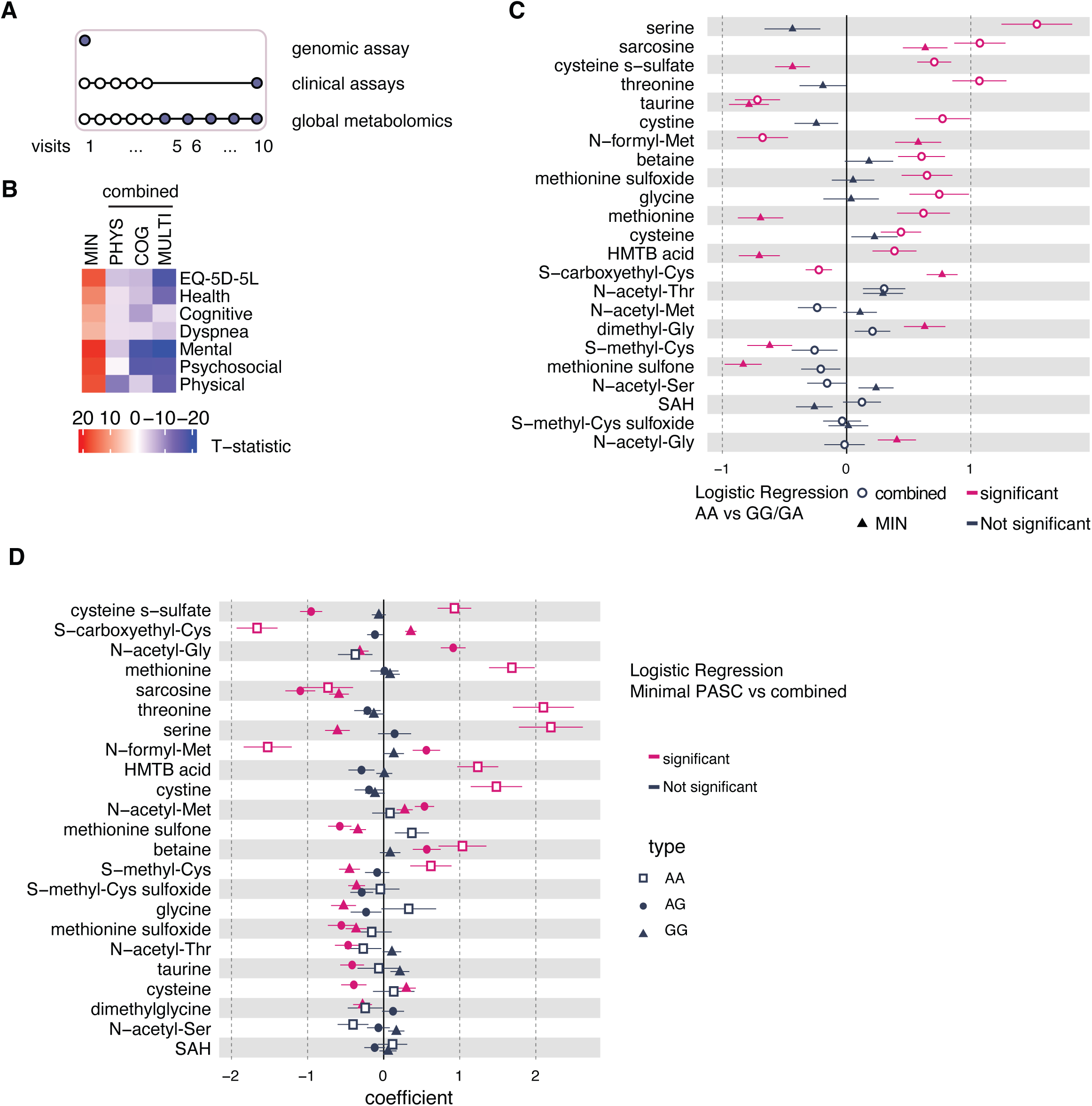
Methionine metabolism alterations during long COVID-19 correlate with *MTHFR* allele status. A A schematic illustrating the study design and employed assays of the IMPACC cohort to generate the subsequent analysis in the figure. Data from genomics, global metabolomics and clinical assays were used. B A heatmap depicting capture of post-acute sequelae of SARS-CoV-2 (PASC), also known as long COVID-19, status in the context of the IMPACC study. Relative deficit for each of four convalescent clusters across seven participant-reported outcomes^5^ is depicted: EQ-5D-5L, Health Recovery Score (Health), PROMIS Cognitive Function Score (Cognitive), PROMIS Psychosocial Illness Impact Positive Score (Psychosocial), PROMIS Global Mental Health Score (Mental), PROMIS Dyspnea Score (Dyspnea), and PROMIS Physical Function Score (Physical). The color coding denotes a t-statistic comparing the within-cluster mean to the remaining sample mean, with t = 0 denoting the overall sample mean and negative values denoting a deficit. The 4 clusters were: minimal deficit (MIN); physical predominant deficit (PHY); mental/cognitive predominant deficit (COG); and multidomain deficit (MLT). PHY, COG and MTL were combined into a single measurement of health deficit, “combined”, to generate the subsequent analysis in the figure. PROMIS Patient-Reported Outcomes Measurement Information System. C Logistic regression analysis for minimal PASC (triangle), and combined PASC (open circle) for indicated metabolites from global metabolomics and either wild type and heterozygous *MTHFR* alleles (GG or GA) or hypomorph alleles (AA), adjusted for sex, age quantile, and BMI. The analysis included samples from visit 1 of trajectory groups 1-4. Significantly different metabolites (p < 0.01, coefficient > 0) are in magenta. AA n = 676; GG/GA n = 3406; D Logistic regression analysis for wild type (GG, triangle symbols), heterozygous (GA, circle) or hypomorph (AA, open square) for indicated metabolites from global metabolomics predicting either minimal or combined PASC clinical outcome, adjusted for sex, age quantile, and BMI. The analysis included samples from visit 1 of trajectory groups 1-4. Significantly different metabolites (p < 0.01, coefficient > 0) are indicated with a circle. MIN n = 728; combined n = 676;

Previously, longitudinal PROs collected during the convalescent period were modelled using latent class mixed models (LCMMs) to identify groups of participants with similar longitudinal patterns^5^. PROs were collected using 7 self-reported health survey questionnaires: EQ-5D-5L (European Quality of Life 5 Dimensions 5 Level Version) score (EQ-5D-5L), Health Recovery Score, PROMIS Cognitive Function Score (Cognitive), PROMIS Psychosocial Illness Impact Positive Score (Psychosocial), PROMIS Global Mental Health Score (Mental), PROMIS Dyspnea Score (Dyspnea), and PROMIS Physical Function Score (Physical). The EQ-5D-5L survey assesses the participant’s health state through 5 questions regarding mobility, self-care, usual activities, pain/discomfort, and anxiety/depression, in which participants rate their symptoms for each category on a scale of 1-none to 5-extreme. The Health Recovery Score is a percentage rating from the participant, comparing their health after they were discharged to their health before their COVID-19 infection and hospitalization. The 5 remaining surveys were from the Common Fund’s Patient-Reported Outcomes Measurement Information System (PROMIS). Using five clustering methods (Ward, McQuitty, Average, PAM, and Complete) and the Gower distance matrix, the Ward algorithm with 6 clusters was selected as the optimal model for our data based on four fitting statistics (Dunn index, average silhouette width, ratio of within to between sum of squares, and within-cluster sum of squares)^5^. To estimate the strength of association of each PRO with each PASC cluster, t-statistics comparing mean PRO scores of one PASC group across the remaining PASC groups were calculated, which identified no associated deficit among 3 Ward clusters, which were then combined into a singular cluster defined as ‘minimal’. This resulted in four PASC groups which are labeled based on their associations with specific PROs: minimal, physical, mental/cognitive, and multidomain (Figure 4B). The t-statistics were inversely coded so that a negative value indicates a worse self-reported health condition. For patients in the “minimal outcome” group a large positive t-statistic score is observed in all PASC categories – indicating lower mean health deficit scores (where lower score indicates better health) in the minimal PASC category compared to the mean deficits in the other PASC categories. The remaining three PASC categories were named based on the deficits with more negative t-statistics, and therefore greater deficits, for that category. In our analysis, we combined the three deficit categories (physical, mental/cognitive, and multidomain) into a single one, referred to as “combined” from here on (Figure 4B).

We performed logistic regression analysis of metabolite abundance at Visit 1 for metabolites relevant for the one-carbon metabolism pathway comparing the wild type (GG) and heterozygous (GA) alleles of *MTHFR* to the homozygous hypomorph (AA) allele for either the minimal or combined PASC outcomes (Figure 4C). For patients experiencing PASC (“combined PASC”), more metabolites were significantly different if they carried the AA allele. Interestingly, significant changes of some metabolites, such as cysteine s−sulfate, methionine or HMTB acid, vary in opposite direction in the minimal compared to combined PASC, suggesting that levels of these metabolites could be adequate biomarkers or to inform on mechanisms of the convalescent disease. This analysis implied that the genetic status of the patient for the *MTHFR* allele is predictive of the metabolic changes observed as early as in Visit 1 for minimal and combined PASC. Therefore, we asked whether stratification of the IMPACC cohort by *MTHFR* allele status can be predictive of metabolic changes when comparing PASC outcome (minimal to combined). Indeed, the analysis indicated that patients with the AA allele exhibited greater differences in metabolite levels as early as in Visit 1 when comparing the PASC outcome of these patients (minimal to combined PASC) (Figure 4D). This suggests that patients with the *MTHFR* AA allele are more likely to suffer from significant metabolic consequences of the disease. We also looked at individual metabolites and noted several significant differences between the metabolite levels in patients with the different alleles depending on PASC severity (Figure S5A). For example, methionine-sulfone levels at Visit 1 were lower in individuals with the AA allele in the combined PASC category (AA vs GG W=1254, p=0.03; AA vs AG W=964, p=0.019) but not in patients who reported minimal deficits. Together our data suggest that *MTHFR* allele status is a genetic predisposition that feeds into early metabolic changes, already observed at Visit 1, that can predict convalescence COVID-19 outcome. These findings could be valuable for personalized medicine approaches, where understanding the genetic background may inform risk stratification, prognosis, or tailored therapeutic interventions.

## Discussion

COVID-19 continues to place a significant burden on human health and the healthcare system, emphasizing the need to predict which patients may require intensive care, and which are at higher risk for developing long COVID-19. In this study, we identified that intermediates of the methionine cycle are significantly altered during disease progression and vary with disease severity. By focusing on changes at the time of hospitalization, we demonstrated that the relative concentrations of specific methionine cycle metabolites correlate with disease severity. We found that the common genetic variant *MTHFR* C677T does not fully account for these correlations. Importantly, incorporating *MTHFR* allele status and a predictive model based on significant changes in methionine-related metabolites resulted in an informative predictive model for both disease severity and for the development of post-acute sequelae of SARS-CoV-2. These findings highlight the potential synergy between genetic and metabolic markers for patient stratification, for informing the need for more aggressive treatment and closer patient observation for patients at risk.

Our findings are actionable: both genetic testing of the gene *MTHFR* and plasma metabolomics can be applied upon hospitalization. Because our data suggest that these parameters are informative already at the first visit in the hospital, stratifying patients by this risk factor can be done early on and inform on following treatment and medical recommendations. personalized approach, that will combine genetic and metabolite information, can be used to build predictive models for disease severity and PASC. However, we emphasize that further studies are needed to validate these results and provide deeper mechanistic insights before implementation in a clinical setting.

Mechanistically, it is not understood if perturbed methionine metabolism plays a causal role in the disease trajectory, or only reflects it. A previous study reported that the *MTHFR* gene polymorphism C677T is significantly associated with the severity of COVID-19 (p = 0.035, n=226)^38^. However, in our study, the genetic status of *MTHFR* allele was not sufficient for prediction of severity, and only when genetic and metabolic data were combined, an informative prediction model for disease severity could be structured. We hypothesize that the genetic status of *MTHFR* provides a predisposition, that when combined with other factors results in pathological manifestation that is related (either as a cause or a result) to COVID-19 severity. The factors that can change the manifestation of the metabolic state include environmental, dietary, immunological, infectious disease history, and possibly others^39, 40^. These factors contribute to variations in the abundance of key methionine metabolism metabolites, as reflected in our metabolite profiling. This hypothesis is a “two-hit” hypothesis, where *MTHFR* allele status is the first, pre-existing, hit, and the metabolite profile at time of hospitalization is the second hit. While the factors that dictate metabolic profile at time of hospitalization are not yet defined, nonetheless, this profile can potentially be used to inform precision care and improve disease outcome.

Both prolonged hospitalization and greater disease severity have been linked to an increased risk of developing PASC (39333538, 35952496, 37880596). Disruptions in DNA methylation have also been associated with PASC. Distinct differentially methylated regions (DMRs) have been identified in PASC patients, which not only distinguish PASC cases from controls but also stratify PASC severity, suggesting their potential as biomarkers (39024897). Our findings fit into this picture and indicate that *MTHFR* genetic variation can influence PASC symptoms through its impact on methionine cycle metabolites, such as homocysteine. Elevated homocysteine levels, linked to reduced MTHFR activity, contribute to endothelial dysfunction and systemic inflammation^41, 42^, which are central to PASC pathophysiology. These results highlight the intersection of epigenetic changes and metabolic dysregulation in PASC, underscoring the need for further research to explore their persistence and role in disease progression. This approach may inform strategies to mitigate long-term complications.

The IMPACC cohort offers significant advantages, including its large and diverse population drawn from multiple geographically dispersed hospitals across the U.S. and its longitudinal design, which tracked participants from acute COVID-19 hospitalization through one year of post-acute recovery. The study’s detailed clinical and biological phenotyping, innovative assays, and standardized data analysis pipelines and high quality data management provided a robust framework for investigating disease mechanisms. Notably, we successfully integrated metabolomics data from two distinct platforms, demonstrating strong agreement between them.

However, the study was limited by the absence of assays to explore methylation or other epigenetic effects, which are likely influenced by disruptions in the methylation cycle and 1CM. Despite our sample size of >1,000 patients, we are still underpowered in genomics data, and other genomics datasets are indeed much bigger. Yet, our unique multi-assayed IMPACC database allowed us to combine the genomics data with metabolomics data for empowerment of our prediction model. Future research can build on these findings to investigate the mechanisms underlying 1CM dysregulation and its impact on COVID-19 severity and PASC phenotypes.

## Data Availability

Data used in this study is available at ImmPort Shared Data under the accession number SDY1760 and in the NLM's Database of Genotypes and Phenotypes (dbGaP) under the accession number phs002686.v1.p1. All code is deposited on Bitbucket (https://bitbucket.org/kleinstein/impacc-public-code/chronic_viruses).

https://www.ncbi.nlm.nih.gov/projects/gap/cgi-bin/study.cgi?study_id=phs002686.v1.p1

## Acknowledgement & Funding

We thank the study participants and local clinical staff across the participating hospitals for making this study possible. We thank all members of the Kanarek lab for their contribution to this work. This study was funded in part by NIH/NIAID Human Immunology Project Consortium U19 award AI118608 (PI Ofer Levy). BCH BTREC Fund (N.K.). N.K. is a Pew Biomedical Scholar.

We thank the participants of the study for their voluntary enrollment and contribution of samples for this work. See the supplement for details on the IMPACC Network. We acknowledge the assistance of the following individuals: Sanya Thomas, Mitchell Cooney, Shun Rao, Sofia Vignolo, and Elena Morrocchi (all from the CDCC); Arash Naeim, Marianne Bernardo, Sarahmay Sanchez, Shannon Intluxay, Clara Magyar, Jenny Brook, Estefania Ramires-Sanchez, Megan Llamas, Claudia Perdomo, Clara E. Magyar, and Jennifer A. Fulcher (all from the David Geffen School of Medicine at UCLA); members of the UCLA Center for Pathology Research Services and the Pathology Research Portal; M. Catherine Muenker, Dimitri Duvilaire, Maxine Kuang, William Ruff, Khadir Raddassi, Denise Shepherd, Haowei Wang, Omkar Chaudhary, Syim Salahuddin, John Fournier, Michael Rainone, and Maxine Kuang (all from the Yale School of Medicine). We thank the leadership of Boston Children’s Hospital including Drs. Wendy Chung, Gary Fleisher and Kevin Churchwell for their support for the Precision Vaccines Program. Dr. Augustine’s and Becker’s co-authorship of this report does not necessarily represent the official views of the National Institute of Allergy and Infectious Diseases, the National Institutes of Health or any other agency of the United States Government.

We thank the participants of the study for their voluntary enrollment and contribution of samples for this work. See the supplement for details on the IMPACC Network. We acknowledge the assistance of the following individuals: Sanya Thomas, Mitchell Cooney, Shun Rao, Sofia Vignolo, and Elena Morrocchi (all from the CDCC); Arash Naeim, Marianne Bernardo, Sarahmay Sanchez, Shannon Intluxay, Clara Magyar, Jenny Brook, Estefania Ramires-Sanchez, Megan Llamas, Claudia Perdomo, Clara E. Magyar, and Jennifer A. Fulcher (all from the David Geffen School of Medicine at UCLA); members of the UCLA Center for Pathology Research Services and the Pathology Research Portal; M. Catherine Muenker, Dimitri Duvilaire, Maxine Kuang, William Ruff, Khadir Raddassi, Denise Shepherd, Haowei Wang, Omkar Chaudhary, Syim Salahuddin, John Fournier, Michael Rainone, and Maxine Kuang (all from the Yale School of Medicine). We thank the leadership of Boston Children’s Hospital including Drs. Wendy Chung, Gary Fleisher and Kevin Churchwell for their support for the Precision Vaccines Program.

## Materials and Methods

### IMPACC study cohort

The IMPACC cohort enrolled 1,164 unvaccinated patients hospitalized with SARS-CoV-2 infection (confirmed by RT-PCR) from 20 hospitals linked to geographically diverse academic institutions across the U.S. between May 5th, 2020 and March 19th, 2021^22, 43^. Details regarding the complete study design, clinical and biological sample collection, and participants’ demographics have been previously outlined^37–39^. In brief, detailed clinical evaluations and blood samples were gathered within 72 hours of hospitalization (visit 1), on days 4, 7, 14, 21, and 28, 90, 180, and 360 following hospital admission (visits 2–6, respectively) for the acute phase, and at 3-, 6-, 9-, and 12-months post-discharge for the convalescent phase^43^.

### Ethics

NIAID staff conferred with the Department of Health and Human Services Office for Human Research Protections (OHRP) regarding potential applicability of the public health surveillance exception [45CFR46.102 (l) (2)] to the IMPACC study protocol. OHRP concurred that the study satisfied criteria for the public health surveillance exception, and the IMPACC study team sent the study protocol and participant information sheet for review and assessment to institutional review boards (IRBs) at participating institutions. Twelve institutions elected to conduct the study as public health surveillance, while three sites with prior IRB-approved biobanking protocols elected to integrate and conduct IMPACC under their institutional protocols (University of Texas at Austin, IRB 2020-04-0117; University of California San Francisco, IRB 20-30497; Case Western reserve university, IRB STUDY20200573) with informed consent requirements. Participants enrolled under the public health surveillance exclusion were provided information sheets describing the study, samples to be collected, and plans for data de-identification and use. Those that requested not to participate after reviewing the information sheet were not enrolled. Participants did not receive compensation for study participation while hospitalized and subsequently were offered compensation during outpatient follow-up.

### Host genotyping

DNA was extracted and samples were genotyped on the Illumina Global Diversity Array as previously described^23^. For this analysis, we extracted the MTHFR rs1801133 (C677T) and rs1801131 (A1298AC) polymorphism.

### Clinical outcome variables

Group-based trajectory modeling, a likelihood-based approach commonly used to group time series of clinical data and previously described^43^, was used to cluster longitudinal measures of the World Health Organization (WHO) 7-point severity ordinal scale into five trajectory groups (TG). Participants with mild disease were defined as those with TG 1-3, and participants with severe disease as those with TG 4-5. TG5 represents all fatal cases within 28-days of admission. Additionally, patients with confirmed mortality at any time during the study were identified.

### Randomization of samples for metabolomics

To mitigate potential batching effects, a randomization procedure was developed to help ensure that longitudinal samples from the same participants were run on the same plates and were randomly distributed across the plates. We stratified this randomization by disease severity (mild/moderate versus severe) and age (younger versus older) with the representation of these strata across plates. In addition, we verified that race, ethnicity, gender, and site were well represented across the plates.

Randomization was applied for both global and targeted metabolomics.

### Global metabolomics

Plasma metabolite profiling was conducted by Metabolon using their proprietary standards. Samples were randomized into batches and prepared with Metabolon’s solvent extraction method, adding recovery standards for quality control. Proteins were precipitated with methanol, shaken, and centrifuged. The supernatants were split into five fractions for analysis with UPLC-MS/MS using various ionization modes, including reverse-phase and HILIC. Instruments included a Waters ACQUITY UPLC and a Thermo Scientific Q-Exactive mass spectrometer.

All identified metabolites met Level 1 metabolite identification standards as established by the Chemical Analysis Working Group of the Metabolomics Standards Initiative (Members et al., 2007; Spicer et al., 2017; Sumner et al., 2007). Orthogonal analytical techniques were applied to confirm the identity of key metabolites. Only metabolites with confirmed accurate mass, retention index, and chemical composition were reported.

The raw data consist of metabolites, each annotated to one of nine super pathways: Amino Acid, Carbohydrate, Cofactors and Vitamins, Energy, Nucleotide, Lipid, Peptide, Xenobiotics, and Partially Characterized Molecules. Metabolite levels were measured using LC-MS peak areas, which are proportional to the concentration of each feature.

For quality control, missing values were imputed using half the minimum detected level for each metabolite. Metabolites with zero interquartile range were excluded from further analysis^23^. All features underwent log-transformation, normalization, and Pareto scaling to minimize variation in fold-change differences.

### Preparation of plasma samples for metabolomics

Plasma samples were collected as described in the IMPACC publication^22^. Originally, 500 µL aliquots were flash frozen and stored at -80 °C in our facility. Aliquots were thawed on ice and re-aliquoted into a new tube at 5 µL to be used for polar metabolomics and folate forms analysis. At this point mock samples were also prepared (empty tubes, left open for the same duration as plasma tubes and handles in parallel) and an aliquot (5 µL) of a universal plasma donor (UPD) mix was thawed. We planned for 3 separate extraction and analysis batches and samples were thawed and aliquoted in subsequent weeks prior to analysis. Per batch, plasma metabolites from a 5 µL aliquot were extracted in 200 µL extraction buffer (80% Methanol, 25 mM Ammonium Acetate and 2.5 mM Na-Ascorbate prepared in LC-MS water, supplemented with isotopically labeled amino acid standards [Cambridge Isotope Laboratories, MSK-A2-1.2], aminopterin, and reduced glutathione standard [Cambridge Isotope Laboratories, CNLM-6245-10]). Samples were vortexed for 10 sec, then centrifuged for 10 minutes at 18,000g to pellet cell debris. The supernatant was transferred into a new tube and dried on ice using a liquid nitrogen dryer.

Metabolites were reconstituted in 100 µL water supplemented with QReSS (at 1/1000, isotopically labeled standards [Cambridge Isotope Laboratories, MSK-QRESS-KIT]) and immediately queued for analysis of folate forms. After the folate run was completed (2 days for about 100 samples per batch) samples were queued for analysis of polar metabolites. This ensured that the more unstable folates will be analyzed first.

### Detection of folate forms from plasma

For targeted metabolomics of folate forms, 7 µL of reconstituted plasma metabolites (equivalent to 0.62 µL plasma) were injected into an Ascentis Express C18 HPLC column (2.7 um x 15 um x 2.1 mm; Sigma Aldrich). The column oven and autosampler tray were held at 30 °C and 4 °C, respectively. The following conditions were used to achieve chromatographic separation: buffer A was 0.1% formic acid; buffer B was acetonitrile with 0.1% formic acid. The chromatographic gradient was run at a flow rate of 0.250 ml min−1 as follows: 0–5 min: gradient was held at 5% B; 5–10 min: linear gradient of 5% to 36% B; 10.1–14.0 min: linear gradient from 36– 95% B; 14.1–18.0 min: gradient was returned to 5% B. The mass spectrometer was operated in full-scan, positive ionization mode using three narrow-range scans: 438–450 m/z; 452–462 m/z; and 470–478 m/z, with the resolution set at 70,000, the AGC target at 10and the maximum injection time of 150 ms. HESI settings were: sheath gas flow rate: 40 psi; Aux gas flow rate: 10 psi; Sweep gas: 0 psi; Spray voltage: 2.8 kV (neg) 3.5 kV (pos); Capillary temperature 300 °C; S-lens RF level 50 (a.u.); Aux gas heater temperature: 350 °C.

### Targeted polar metabolomics by LC-MS

For targeted polar metabolomics, 1 µL reconstituted plasma metabolites (equivalent to 0.05 µL plasma) was injected into a ZIC-pHILIC 150 x 2.1 mm (5 um particle size) column (EMD Millipore). operated on a Vanquish™ Flex UHPLC Systems (Thermo Fisher Scientific, San Jose, CA, USA). Chromatographic separation was achieved using the following conditions: buffer A was acetonitrile; buffer B was 20 mM ammonium carbonate, 0.1% ammonium hydroxide in water; resulting pH is around 9 without pH adjustment. Gradient conditions we used were: 0–20 min: linear gradient from 20% to 80% B; 20–20.5 min: from 80% to 20% B; 20.5–28 min: hold at 20% B at 150 uL/min flow rate. The column oven and autosampler tray were held at 25 °C and 4 °C, respectively. MS data acquisition was performed using a QExactive benchtop orbitrap mass spectrometer equipped with an Ion Max source and a HESI II probe (Thermo Fisher Scientific, San Jose, CA, USA) and was performed in positive and negative ionization mode in a range of m/z = 70–1000, with the resolution set at 70,000, the AGC target at 1 × 10^6^, and the maximum injection time (Max IT) at 20 msec. HESI settings were: Sheath gas flow rate: 35 psi. Aux gas flow rate: 8 psi. Sweep gas flow rate: 1 psi. Spray voltage 3.5 kV (pos); 2.8 kV (neg). Capillary temperature: 320 °C. S-lens RF level: 50 (a.u.). Aux gas heater temperature: 350 °C.

### Data Analysis for targeted metabolomics

Relative quantification of polar metabolites and folate forms was performed with TraceFinder 5.1 (Thermo Fisher Scientific, Waltham, MA, USA) using a 7 ppm mass tolerance and referencing an in-house library of chemical standards (220 standards, 7 folate forms and 37 internal standards). Pooled samples and fractional dilutions were prepared as quality controls and injected at the beginning and end of each run. In addition, pooled samples were interspersed throughout the run to control for technical drift in signal quality as well as to serve to assess the coefficient of variability (CV) for each metabolite. In addition to pooled samples, we interspersed injections of the UPD samples prepared in parallel for each batch to assist in batch normalization and data merging. Data from TraceFinder was further consolidated and normalized manually in Excel. We further followed a normalization strategy originally described in Wulff et al.^25^ to synchronize between Metabolon normalized data and our targeted approach. Briefly, relying on the pooled sample re-injections and dilutions we assigned respectively a CV (coefficient of variation) and an RSQ (r-squared) values to each metabolite per batch. Then performed a filtering step and retained only metabolites for which CV<30% and RSQ>0.95 in at least two of the three batches. Finally, for each metabolite values were mean centered across samples and batches. This strategy effectively minimized batch effects and allowed for relative comparisons. Finally, data was log-transformed and Pareto scaled for all multivariate statistics analysis. Individual graphs were plotted in GraphPad (Prism) while pathway analysis, and heatmaps were performed on the MetaboAnalyst 6.0 platform. For the analysis of folate forms similar strategy was followed and post batch normalization folate forms were integrated within polar metabolites dataset. A single outlier was present in batch 1, for which the sample tube was found to contain no plasma. Despite this the tube was processed as a mock sample not to interfere with the strict randomization strategy.

## #The IMPACC Network

**Clinical and Data Coordinating Center (CDCC), Precision Vaccines Program, Boston Children’s Hospital, Harvard Medical School, Boston, MA 02115, USA:**

Al Ozonoff, Joann Diray-Arce, Jing Chen, Alvin T. Kho, Carly E. Milliren, Annmarie Hoch, Ana C. Chang, Kerry McEnaney, Caitlin Syphurs, Brenda Barton, Claudia Lentucci, Maimouna D. Murphy, Mehmet Saluvan, Tanzia Shaheen, Shanshan Liu, Marisa Albert, Arash Nemati Hayati, Robert Bryant, James Abraham, Mitchell Cooney, Meagan Karoly

**Benaroya Research Institute, University of Washington, Seattle, WA 98101, USA:** Matthew C. Altman, Naresh Doni Jayavelu, Scott Presnell, Bernard Kohr, Tomasz Jancsyk, Azlann Arnett

**La Jolla Institute for Immunology, La Jolla, CA 92037, USA:**

Bjoern Peters, James A. Overton, Randi Vita, Kerstin Westendorf

**Knocean Inc. Toronto, ON M6P 2T3, Canada:**

James A. Overton

**Precision Vaccines Program, Boston Children’s Hospital, Harvard Medical School, Boston, MA 02115, USA:**

Ofer Levy, Hanno Steen, Patrick van Zalm, Benoit Fatou, Kinga K. Smolen, Arthur Viode, Simon van Haren, Meenakshi Jha, David Stevenson, Athena N. Nguyen, Alec L. Plotkin, Sanya Thomas, Boryana Petrova, Naama Kanarek

**Brigham and Women’s Hospital, Harvard Medical School, Boston, MA 02115, USA:** Lindsey R. Baden, Kevin Mendez, Jessica Lasky-Su, Alexandra Tong, Rebecca Rooks, Michael Desjardins, Amy C. Sherman, Stephen R. Walsh, Xhoi Mitre, Jessica Cauley, Xiaofang Li, Bethany Evans, Christina Montesano, Jose Humberto Licona, Jonathan Krauss, Nicholas C. Issa, Jun Bai Park Chang, Natalie Izaguirre

**Metabolon Inc, Morrisville, NC 27560, USA:**

Scott R. Hutton, Greg Michelotti, Kari Wong

**Prevention of Organ Failure (PROOF) Centre of Excellence, University of British Columbia, Vancouver, BC V6T 1Z3, Canada:**

Scott J. Tebbutt, Casey P. Shannon

**Case Western Reserve University and University Hospitals of Cleveland, Cleveland, OH 44106, USA:**

Rafick-Pierre Sekaly, Slim Fourati, Grace A. McComsey, Paul Harris, Scott Sieg, George Yendewa, Mary Consolo, Heather Tribout, Susan Pereira Ribeiro

**Drexel University, Tower Health Hospital, Philadelphia, PA 19104, USA:**

Charles B. Cairns, Elias K. Haddad, Michele A. Kutzler, Mariana Bernui, Gina Cusimano, Jennifer Connors, Kyra Woloszczuk, David Joyner, Carolyn Edwards, Edward Lee, Edward Lin, Nataliya Melnyk, Debra L. Powell, James N. Kim, I. Michael Goonewardene, Brent Simmons, Cecilia M. Smith, Mark Martens, Brett Croen, Nicholas C. Semenza, Mathew R. Bell, Sara Furukawa, Renee McLin, George P. Tegos, Brandon Rogowski, Nathan Mege, Kristen Ulring, Pam Schearer, Judie Sheidy, Crystal Nagle

**MyOwnMed Inc., Bethesda, MD 20817, USA:**

Vicki Seyfert-Margolis

**Emory School of Medicine, Atlanta, GA 30322, USA:**

Nadine Rouphael, Steven E. Bosinger, Arun K. Boddapati, Greg K. Tharp, Kathryn L. Pellegrini, Brandi Johnson, Bernadine Panganiban, Christopher Huerta, Evan J. Anderson, Hady Samaha, Jonathan E. Sevransky, Laurel Bristow, Elizabeth Beagle, David Cowan, Sydney Hamilton, Thomas Hodder, Amer Bechnak, Andrew Cheng, Aneesh Mehta, Caroline R. Ciric, Christine Spainhour, Erin Carter, Erin M. Scherer, Jacob Usher, Kieffer Hellmeister, Laila Hussaini, Lauren Hewitt, Nina Mcnair, Susan Pereira Ribeiro, Sonia Wimalasena

**Icahn School of Medicine at Mount Sinai, New York, NY 10029, USA:**

Ana Fernandez-Sesma, Viviana Simon, Florian Krammer, Harm Van Bakel, Seunghee Kim-Schulze, Ana Silvia Gonzalez-Reiche, Jingjing Qi, Brian Lee, Juan Manuel Carreño, Gagandeep Singh, Ariel Raskin, Johnstone Tcheou, Zain Khalil, Adriana van de Guchte, Keith Farrugia, Zenab Khan, Geoffrey Kelly, Komal Srivastava, Lily Q. Eaker, Maria C. Bermúdez-González, Lubbertus C.F. Mulder, Katherine F. Beach, Miti Saksena, Deena Altman, Erna Kojic, Levy A. Sominsky, Arman Azad, Dominika Bielak, Hisaaki Kawabata, Temima Yellin, Miriam Fried, Leeba Sullivan, Sara Morris, Giulio Kleiner, Daniel Stadlbauer, Jayeeta Dutta, Hui Xie, Manishkumar Patel, Kai Nie, Brian Monahan

**Immunai Inc., New York, NY 10016, USA:**

Adeeb Rahman

**Oregon Health & Science University, Portland, OR 97239, USA:**

William B. Messer, Catherine L. Hough, Sarah A.R. Siegel, Peter E. Sullivan, Zhengchun Lu, Amanda E. Brunton, Matthew Strand, Zoe L. Lyski, Felicity J. Coulter, Courtney Micheletti

**Stanford University School of Medicine, Palo Alto, CA 94305, USA:**

Holden Maecker, Bali Pulendran, Kari C. Nadeau, Yael Rosenberg-Hasson, Michael Leipold, Natalia Sigal, Angela Rogers, Andrea Fernandes, Monali Manohar, Evan Do, Iris Chang, Alexandra S. Lee, Catherine Blish, Henna Naz Din, Jonasel Roque, Linda N. Geng, Maja Artandi, Mark M. Davis, Neera Ahuja, Samuel S. Yang, Sharon Chinthrajah, Thomas Hagan, Tyson H. Holmes, Koji Abe

**David Geffen School of Medicine at the University of California Los Angeles, Los Angeles CA 90095, USA:**

Elaine F. Reed, Joanna Schaenman, Ramin Salehi-Rad, Adreanne M. Rivera, Harry C. Pickering, Subha Sen, David Elashoff, Dawn C. Ward, Jenny Brook, Estefania Ramires-Sanchez, Megan Llamas, Claudia Perdomo, Clara E. Magyar, Jennifer Fulcher

**University of California San Francisco, San Francisco, CA 94115, USA:**

David J. Erle, Carolyn S. Calfee, Carolyn M. Hendrickson, Kirsten N. Kangelaris, Viet Nguyen, Deanna Lee, Suzanna Chak, Rajani Ghale, Ana Gonzalez, Alejandra Jauregui, Carolyn Leroux, Luz Torres Altamirano, Ahmad Sadeed Rashid, Andrew Willmore, Prescott G. Woodruff, Matthew F. Krummel, Sidney Carrillo, Alyssa Ward, Charles R. Langelier, Ravi Patel, Michael Wilson, Ravi Dandekar, Bonny Alvarenga, Jayant Rajan, Walter Eckalbar, Andrew W. Schroeder, Gabriela K. Fragiadakis, Alexandra Tsitsiklis, Eran Mick, Yanedth Sanchez Guerrero, Christina Love, Lenka Maliskova, Michael Adkisson, Aleksandra Leligdowicz, Alexander Beagle, Arjun Rao, Austin Sigman, Bushra Samad, Cindy Curiel, Cole Shaw, Gayelan Tietje-Ulrich, Jeff Milush, Jonathan Singer, Joshua J. Vasquez, Kevin Tang, Legna Betancourt, Lekshmi Santhosh, Logan Pierce, Maria Tecero Paz, Michael Matthay, Neeta Thakur, Nicklaus Rodriguez, Nicole Sutter, Norman Jones, Pratik Sinha, Priya Prasad, Raphael Lota, Saurabh Asthana, Sharvari Bhide, Tasha Lea, Yumiko Abe-Jones

**Yale School of Medicine, New Haven, CT 06510, USA:**

David A. Hafler, Ruth R. Montgomery, Albert C. Shaw, Steven H. Kleinstein, Jeremy P. Gygi, Dylan Duchen, Shrikant Pawar, Anna Konstorum, Ernie Chen, Chris Cotsapas, Xiaomei Wang, Charles Dela Cruz, Akiko Iwasaki, Subhasis Mohanty, Allison Nelson, Yujiao Zhao, Shelli Farhadian, Hiromitsu Asashima, Omkar Chaudhary, Andreas Coppi, John Fournier, M. Catherine Muenker, Khadir Raddassi, Michael Rainone, William Ruff, Syim Salahuddin, WadeL. Shulz, Pavithra Vijayakumar, Haowei Wang, Esio Wunder Jr., H. Patrick Young, Albert I. Ko, Gisela Gabernet

**Yale School of Public Health, New Haven, CT 06510, USA:**

Denise Esserman, Leying Guan, Anderson Brito, Jessica Rothman, Nathan D. Grubaugh, Kexin Wang, Leqi Xu

**Baylor College of Medicine and the Center for Translational Research on Inflammatory Diseases, Houston, TX 77030, USA:**

David B. Corry, Farrah Kheradmand, Li-Zhen Song, Ebony Nelson

**Oklahoma University Health Sciences Center, Oklahoma City, OK 73104, USA:**

Jordan P. Metcalf, Nelson I. Agudelo Higuita, Lauren A. Sinko, J. Leland Booth, Douglas A. Drevets, Brent R. Brown

**University of Arizona, Tucson AZ 85721, USA:**

Monica Kraft, Chris Bime, Jarrod Mosier, Heidi Erickson, Ron Schunk, Hiroki Kimura, Michelle Conway, Dave Francisco, Allyson Molzahn, Connie Cathleen Wilson, Ron Schunk, Trina Hughes, Bianca Sierra

**University of Florida, Gainesville, FL 32611, USA:**

Mark A. Atkinson, Scott C. Brakenridge, Ricardo F. Ungaro, Brittany Roth Manning, Lyle Moldawer

**University of Florida, Jacksonville, FL 32218, USA:**

Jordan Oberhaus, Faheem W. Guirgis

**University of South Florida, Tampa FL 33620, USA:**

Brittney Borresen, Matthew L. Anderson

**The University of Texas at Austin, Austin, TX 78712, USA:**

Lauren I. R. Ehrlich, Esther Melamed, Cole Maguire, Dennis Wylie, Justin F. Rousseau, Kerin C. Hurley, Janelle N. Geltman, Nadia Siles, Jacob E. Rogers, Pablo Guaman Tipan

**IMPACC Network Competing Interests**

The Icahn School of Medicine at Mount Sinai has filed patent applications related to SARS-CoV-2 serological assays, NDV-based SARS-CoV-2 vaccines, influenza virus vaccines, and therapeutics, listing Florian Krammer and Viviana Simon as co-inventors. Mount Sinai has spun out Kantaro to market SARS-CoV-2 serological tests and Castlevax to develop SARS-CoV-2 vaccines, with Florian Krammer as co-founder and scientific advisory board member of Castlevax. Florian Krammer has consulted for Merck, Curevac, Seqirus, GSK, Pfizer, 3rd Rock Ventures, Sanofi, Gritstone, and Avimex. His laboratory collaborates with Dynavax on influenza vaccine development and with VIR on influenza virus therapeutics development. Ofer Levy is a named inventor on patents held by Boston Children’s Hospital related to vaccine adjuvants and human in vitro platforms that model vaccine action. His laboratory has received research support from GlaxoSmithKline (GSK) and Pfizer and he is a co-founder and advisor to Ovax, Inc., which develops opioid vaccines. Charles Cairns consults for bioMérieux and receives grant funding from the Bill & Melinda Gates Foundation. James A. Overton is a consultant at Knocean Inc.

Jessica Lasky-Su is a scientific advisor for Precion Inc. Scott R. Hutton, Greg Michelloti, and Kari Wong are employees of Metabolon Inc. Vicki Seyfer-Margolis is employed by MyOwnMed. Nadine Rouphael reports grants or contracts with Merck, Sanofi, Pfizer, Vaccine Company, and Immorna. She has served on data safety monitoring boards for Moderna, Sanofi, Seqirus, Pfizer, EMMES, ICON, BARDA, and CyanVan Micron. She has received travel support from Sanofi and Moderna and honoraria from Virology Education and Krog Consulting. Chris Cotsapas is employed by Vesalius Therapeutics. Adeeb Rahman is employed by Immunai Inc. Steven Kleinstein consults for Peraton related to the ImmPort data repository. Nathan Grubaugh consults for Tempus Labs and the National Basketball Association. Akiko Iwasaki consults for 4BIO, Blue Willow Biologics, Revelar Biotherapeutics, RIGImmune, Xanadu Bio, and Paratus Sciences. Monika Kraft receives research funding from NIH, ALA, Sanofi, and AstraZeneca for asthma research. She consults for AstraZeneca, Sanofi, Chiesi, and GSK for severe asthma and is co-founder and CMO of RaeSedo, Inc., developing peptidomimetics for inflammatory lung disease. Esther Melamed receives research funding from Babson Diagnostics, honoraria from the Multiple Sclerosis Association of America, and has served on advisory boards for Genentech, Horizon, Teva, and Viela Bio. Carolyn Calfee receives research funding from NIH, FDA, DOD, Roche-Genentech, and Quantum Leap Healthcare Collaborative and consults for Janssen, Vasomune, Gen1e Life Sciences, NGMBio, and Cellenkos. Wade Schulz has collaborated with the Shenzhen Center for Health Information and the National Center for Cardiovascular Diseases in Beijing, is a technical consultant for Hugo Health, co-founder of Refactor Health, and has received grants from Merck and Regeneron Pharmaceuticals for COVID-19 research. Grace A. McComsey receives research grants from Redhill, Cognivue, Pfizer, and Genentech and consults for Gilead, Merck, and ViiV/GSK. Linda N. Geng receives research funding from Pfizer, Inc., through her institution. Catherine Hough receives research support from NIH and CDC. David Hafler has received research funding from Bristol-Myers Squibb, Novartis, Sanofi, and Genentech and consults for Bayer Pharmaceuticals, Repertoire Inc., Bristol Myers Squibb, Compass Therapeutics, EMD Serono, Genentech, Novartis Pharmaceuticals, and Sanofi Genzyme.

## Funding

NIH (3U01AI167892-03S2, 3U01AI167892-01S2, 5R01AI135803-03, 5U19AI118608-04,

5U19AI128910-04, 4U19AI090023-11, 4U19AI118610-06, R01AI145835-01A1S1,

5U19AI062629-17, 5U19AI057229-17, 5U19AI057229-18, 5U19AI125357-05,

5U19AI128913-03, 3U19AI077439-13, 5U54AI142766-03, 5R01AI104870-07, 3U19AI089992-

09, 3U19AI128913-03, and 5T32DA018926-18); NIAID, NIH (3U19AI1289130,

U19AI128913-04S1, and R01AI122220); NCATS, NIH UM1TR004528 and National Science

Foundation (DMS2310836).

## References

1. Wang J, Goodfellow H, Walker S, Blandford A, Pfeffer P, Hurst JR, Sunkersing D, Bradbury K, Robson C, Henley W, Gomes M. Trajectories of functional limitations, health-related quality of life and societal costs in individuals with long COVID: a population-based longitudinal cohort study. BMJ Open. 2024;14(11):e088538. Epub 20241113. doi: 10.1136/bmjopen-2024-088538. PubMed PMID: 39537389; PMCID: PMC11574431.

2. CDC 2024. Available from: www.cdc.gov/covid/php/surveillance/burden-estimates.html.

3. Groff D, Sun A, Ssentongo AE, Ba DM, Parsons N, Poudel GR, Lekoubou A, Oh JS, Ericson JE, Ssentongo P, Chinchilli VM. Short-term and Long-term Rates of Postacute Sequelae of SARS-CoV-2 Infection: A Systematic Review. JAMA Netw Open. 2021;4(10):e2128568. Epub 20211001. doi: 10.1001/jamanetworkopen.2021.28568. PubMed PMID: 34643720; PMCID: PMC8515212.

4. Admon AJ, Iwashyna TJ, Kamphuis LA, Gundel SJ, Sahetya SK, Peltan ID, Chang SY, Han JH, Vranas KC, Mayer KP, Hope AA, Jolley SE, Caldwell E, Monahan ML, Hauschildt K, Brown SM, Aggarwal NR, Thompson BT, Hough CL. Assessment of Symptom, Disability, and Financial Trajectories in Patients Hospitalized for COVID-19 at 6 Months. JAMA Netw Open. 2023;6(2):e2255795. Epub 20230201. doi: 10.1001/jamanetworkopen.2022.55795. PubMed PMID: 36787143; PMCID: PMC9929698.

5. Ozonoff A, Jayavelu ND, Liu S, Melamed E, Milliren CE, Qi J, Geng LN, McComsey GA, Cairns CB, Baden LR, Schaenman J, Shaw AC, Samaha H, Seyfert-Margolis V, Krammer F, Rosen LB, Steen H, Syphurs C, Dandekar R, Shannon CP, Sekaly RP, Ehrlich LIR, Corry DB, Kheradmand F, Atkinson MA, Brakenridge SC, Higuita NIA, Metcalf JP, Hough CL, Messer WB, Pulendran B, Nadeau KC, Davis MM, Sesma AF, Simon V, van Bakel H, Kim-Schulze S, Hafler DA, Levy O, Kraft M, Bime C, Haddad EK, Calfee CS, Erle DJ, Langelier CR, Eckalbar W, Bosinger SE, Peters B, Kleinstein SH, Reed EF, Augustine AD, Diray-Arce J, Maecker HT, Altman MC, Montgomery RR, Becker PM, Rouphael N. Features of acute COVID-19 associated with post-acute sequelae of SARS-CoV-2 phenotypes: results from the IMPACC study. Nat Commun. 2024;15(1):216. Epub 20240103. doi: 10.1038/s41467-023-44090-5. PubMed PMID: 38172101; PMCID: PMC10764789.

6. Hill EL, Mehta HB, Sharma S, Mane K, Singh SK, Xie C, Cathey E, Loomba J, Russell S, Spratt H, DeWitt PE, Ammar N, Madlock-Brown C, Brown D, McMurry JA, Chute CG, Haendel MA, Moffitt R, Pfaff ER, Bennett TD. Risk factors associated with post-acute sequelae of SARS-CoV-2: an N3C and NIH RECOVER study. BMC Public Health. 2023;23(1):2103. Epub 20231025. doi: 10.1186/s12889-023-16916-w. PubMed PMID: 37880596; PMCID: PMC10601201.

7. Zhang Y, Guo R, Kim SH, Shah H, Zhang S, Liang JH, Fang Y, Gentili M, Leary CNO, Elledge SJ, Hung DT, Mootha VK, Gewurz BE. SARS-CoV-2 hijacks folate and one-carbon metabolism for viral replication. Nat Commun. 2021;12(1):1676. Epub 20210315. doi: 10.1038/s41467-021-21903-z. PubMed PMID: 33723254; PMCID: PMC7960988.

8. Ducker GS, Rabinowitz JD. One-Carbon Metabolism in Health and Disease. Cell Metab. 2017;25(1):27–42. Epub 20160915. doi: 10.1016/j.cmet.2016.08.009. PubMed PMID: 27641100; PMCID: PMC5353360.

9. Wilson PM, Danenberg PV, Johnston PG, Lenz HJ, Ladner RD. Standing the test of time: targeting thymidylate biosynthesis in cancer therapy. Nat Rev Clin Oncol. 2014;11(5):282–98. Epub 20140415. doi: 10.1038/nrclinonc.2014.51. PubMed PMID: 24732946.

10. Perła-Kaján J, Jakubowski H. COVID-19 and One-Carbon Metabolism. Int J Mol Sci. 2022;23(8). Epub 20220410. doi: 10.3390/ijms23084181. PubMed PMID: 35456998; PMCID: PMC9026976.

11. Stegmann KM, Dickmanns A, Gerber S, Nikolova V, Klemke L, Manzini V, Schlösser D, Bierwirth C, Freund J, Sitte M, Lugert R, Salinas G, Meister TL, Pfaender S, Görlich D, Wollnik B, Groß U, Dobbelstein M. The folate antagonist methotrexate diminishes replication of the coronavirus SARS-CoV-2 and enhances the antiviral efficacy of remdesivir in cell culture models. Virus Res. 2021;302:198469. Epub 20210606. doi: 10.1016/j.virusres.2021.198469. PubMed PMID: 34090962; PMCID: PMC8180352.

12. Weisberg I, Tran P, Christensen B, Sibani S, Rozen R. A second genetic polymorphism in methylenetetrahydrofolate reductase (MTHFR) associated with decreased enzyme activity. Mol Genet Metab. 1998;64(3):169–72. doi: 10.1006/mgme.1998.2714. PubMed PMID: 9719624.

13. Liew SC, Gupta ED. Methylenetetrahydrofolate reductase (MTHFR) C677T polymorphism: epidemiology, metabolism and the associated diseases. Eur J Med Genet. 2015;58(1):1–10. Epub 20141104. doi: 10.1016/j.ejmg.2014.10.004. PubMed PMID: 25449138.

14. Karst M, Hollenhorst J, Achenbach J. Life-threatening course in coronavirus disease 2019 (COVID-19): Is there a link to methylenetetrahydrofolic acid reductase (MTHFR) polymorphism and hyperhomocysteinemia? Med Hypotheses. 2020;144:110234. Epub 20200902. doi: 10.1016/j.mehy.2020.110234. PubMed PMID: 33254541; PMCID: PMC7467063.

15. Ponti G, Pastorino L, Manfredini M, Ozben T, Oliva G, Kaleci S, Iannella R, Tomasi A. COVID-19 spreading across world correlates with C677T allele of the methylenetetrahydrofolate reductase (MTHFR) gene prevalence. J Clin Lab Anal. 2021;35(7):e23798. Epub 20210601. doi: 10.1002/jcla.23798. PubMed PMID: 34061414; PMCID: PMC8209953.

16. Kryukov EV, Ivanov AV, Karpov VO, Vasil’evich Aleksandrin V, Dygai AM, Kruglova MP, Kostiuchenko GI, Kazakov SP, Kubatiev AA. Plasma S-Adenosylmethionine Is Associated with Lung Injury in COVID-19. Dis Markers. 2021;2021:7686374. Epub 20211216. doi: 10.1155/2021/7686374. PubMed PMID: 34956420; PMCID: PMC8702356.

17. Karakosta C, Kontou E, Xirou T, Kabanarou SA. Acute Macular Neuroretinopathy Associated With COVID-19 Infection: Is Double Heterozygous Methylenetetrahydrofolate Reductase (MTHFR) Mutation an Underlying Risk Factor? Cureus. 2023;15(2):e34873. Epub 20230211. doi: 10.7759/cureus.34873. PubMed PMID: 36855586; PMCID: PMC9968507.

18. Celestino GG, Amarante MK, Vespero EC, Tavares ER, Yamauchi LM, Candido É D, de Oliveira DBL, Durigon EL, Yamada-Ogatta SF, Faccin-Galhardi LC. Dermatological Manifestations in COVID-19: A Case Study of SARS-CoV-2 Infection in a Genetic Thrombophilic Patient with Mthfr Mutation. Pathogens. 2023;12(3). Epub 20230310. doi: 10.3390/pathogens12030438. PubMed PMID: 36986360; PMCID: PMC10058784.

19. Moness H, Mousa SO, Mousa SO, Adel NM, Ibrahim RA, Hassan EE, Abdelhameed NI, Meshref DA, Abdullah NM. Thrombophilia genetic mutations and their relation to disease severity among patients with COVID-19. PLoS One. 2024;19(3):e0296668. Epub 20240320. doi: 10.1371/journal.pone.0296668. PubMed PMID: 38507367; PMCID: PMC10954113.

20. Balnis J, Madrid A, Drake LA, Vancavage R, Tiwari A, Patel VJ, Ramos RB, Schwarz JJ, Yucel R, Singer HA, Alisch RS, Jaitovich A. Blood DNA methylation in post-acute sequelae of COVID-19 (PASC): a prospective cohort study. EBioMedicine. 2024;106:105251. Epub 20240717. doi: 10.1016/j.ebiom.2024.105251. PubMed PMID: 39024897; PMCID: PMC11286994.

21. Balnis J, Madrid A, Hogan KJ, Drake LA, Adhikari A, Vancavage R, Singer HA, Alisch RS, Jaitovich A. Persistent blood DNA methylation changes one year after SARS-CoV-2 infection. Clin Epigenetics. 2022;14(1):94. Epub 20220723. doi: 10.1186/s13148-022-01313-8. PubMed PMID: 35871090; PMCID: PMC9308917.

22. Immunophenotyping assessment in a COVID-19 cohort (IMPACC): A prospective longitudinal study. Sci Immunol. 2021;6(62). doi: 10.1126/sciimmunol.abf3733. PubMed PMID: 34376480; PMCID: PMC8713959.

23. Diray-Arce J, Fourati S, Doni Jayavelu N, Patel R, Maguire C, Chang AC, Dandekar R, Qi J, Lee BH, van Zalm P, Schroeder A, Chen E, Konstorum A, Brito A, Gygi JP, Kho A, Chen J, Pawar S, Gonzalez-Reiche AS, Hoch A, Milliren CE, Overton JA, Westendorf K, Cairns CB, Rouphael N, Bosinger SE, Kim-Schulze S, Krammer F, Rosen L, Grubaugh ND, van Bakel H, Wilson M, Rajan J, Steen H, Eckalbar W, Cotsapas C, Langelier CR, Levy O, Altman MC, Maecker H, Montgomery RR, Haddad EK, Sekaly RP, Esserman D, Ozonoff A, Becker PM, Augustine AD, Guan L, Peters B, Kleinstein SH. Multi-omic longitudinal study reveals immune correlates of clinical course among hospitalized COVID-19 patients. Cell Rep Med. 2023;4(6):101079. Epub 20230523. doi: 10.1016/j.xcrm.2023.101079. PubMed PMID: 37327781; PMCID: PMC10203880.

24. Gygi JP, Maguire C, Patel RK, Shinde P, Konstorum A, Shannon CP, Xu L, Hoch A, Jayavelu ND, Haddad EK, Reed EF, Kraft M, McComsey GA, Metcalf JP, Ozonoff A, Esserman D, Cairns CB, Rouphael N, Bosinger SE, Kim-Schulze S, Krammer F, Rosen LB, van Bakel H, Wilson M, Eckalbar WL, Maecker HT, Langelier CR, Steen H, Altman MC, Montgomery RR, Levy O, Melamed E, Pulendran B, Diray-Arce J, Smolen KK, Fragiadakis GK, Becker PM, Sekaly RP, Ehrlich LI, Fourati S, Peters B, Kleinstein SH, Guan L. Integrated longitudinal multiomics study identifies immune programs associated with acute COVID-19 severity and mortality. J Clin Invest. 2024;134(9). Epub 20240501. doi: 10.1172/jci176640. PubMed PMID: 38690733; PMCID: PMC11060740.

25. Wulff JaM, M. A Comparison of Various Normalization Methods for LC/MS Metabolomics Data. Advances in Bioscience and Biotechnology. 2018;9:339–51. doi: 10.4236/abb.2018.98022

26. Su Y, Chen D, Yuan D, Lausted C, Choi J, Dai CL, Voillet V, Duvvuri VR, Scherler K, Troisch P, Baloni P, Qin G, Smith B, Kornilov SA, Rostomily C, Xu A, Li J, Dong S, Rothchild A, Zhou J, Murray K, Edmark R, Hong S, Heath JE, Earls J, Zhang R, Xie J, Li S, Roper R, Jones L, Zhou Y, Rowen L, Liu R, Mackay S, O’Mahony DS, Dale CR, Wallick JA, Algren HA, Zager MA, Wei W, Price ND, Huang S, Subramanian N, Wang K, Magis AT, Hadlock JJ, Hood L, Aderem A, Bluestone JA, Lanier LL, Greenberg PD, Gottardo R, Davis MM, Goldman JD, Heath JR. Multi-Omics Resolves a Sharp Disease-State Shift between Mild and Moderate COVID-19. Cell. 2020;183(6):1479–95.e20. Epub 20201028. doi: 10.1016/j.cell.2020.10.037. PubMed PMID: 33171100; PMCID: PMC7598382.

27. Xiao N, Nie M, Pang H, Wang B, Hu J, Meng X, Li K, Ran X, Long Q, Deng H, Chen N, Li S, Tang N, Huang A, Hu Z. Integrated cytokine and metabolite analysis reveals immunometabolic reprogramming in COVID-19 patients with therapeutic implications. Nat Commun. 2021;12(1):1618. Epub 20210312. doi: 10.1038/s41467-021-21907-9. PubMed PMID: 33712622; PMCID: PMC7955129.

28. Páez-Franco JC, Torres-Ruiz J, Sosa-Hernández VA, Cervantes-Díaz R, Romero-Ramírez S, Pérez-Fragoso A, Meza-Sánchez DE, Germán-Acacio JM, Maravillas-Montero JL, Mejía-Domínguez NR, Ponce-de-León A, Ulloa-Aguirre A, Gómez-Martín D, Llorente L. Metabolomics analysis reveals a modified amino acid metabolism that correlates with altered oxygen homeostasis in COVID-19 patients. Sci Rep. 2021;11(1):6350. Epub 20210318. doi: 10.1038/s41598-021-85788-0. PubMed PMID: 33737694; PMCID: PMC7973513.

29. Petrova B, Maynard AG, Wang P, Kanarek N. Regulatory mechanisms of one-carbon metabolism enzymes. J Biol Chem. 2023;299(12):105457. Epub 20231109. doi: 10.1016/j.jbc.2023.105457. PubMed PMID: 37949226; PMCID: PMC10758965.

30. Bailey LB, Gregory JF, 3rd. Polymorphisms of methylenetetrahydrofolate reductase and other enzymes: metabolic significance, risks and impact on folate requirement. J Nutr. 1999;129(5):919–22. doi: 10.1093/jn/129.5.919. PubMed PMID: 10222379.

31. Li Q, Lan Q, Zhang Y, Bassig BA, Holford TR, Leaderer B, Boyle P, Zhu Y, Qin Q, Chanock S, Rothman N, Zheng T. Role of one-carbon metabolizing pathway genes and gene-nutrient interaction in the risk of non-Hodgkin lymphoma. Cancer Causes Control. 2013;24(10):1875–84. Epub 20130803. doi: 10.1007/s10552-013-0264-3. PubMed PMID: 23913011; PMCID: PMC3951097.

32. Nefic H, Mackic-Djurovic M, Eminovic I. The Frequency of the 677C>T and 1298A>C Polymorphisms in the Methylenetetrahydrofolate Reductase (MTHFR) Gene in the Population. Med Arch. 2018;72(3):164–9. doi: 10.5455/medarh.2018.72.164-169. PubMed PMID: 30061759; PMCID: PMC6021155.

33. Heller G, Seshan VE, Moskowitz CS, Gönen M. Inference for the difference in the area under the ROC curve derived from nested binary regression models. Biostatistics. 2016;18(2):260–74. doi: 10.1093/biostatistics/kxw045.

34. Lewis F, Butler A, Gilbert L. A unified approach to model selection using the likelihood ratio test. Methods in Ecology and Evolution. 2011;2(2):155–62. doi: 10.1111/j.2041-210X.2010.00063.x.

35. Vuong QH. Likelihood Ratio Tests for Model Selection and Non-Nested Hypotheses. Econometrica. 1989;57(2):307–33. doi: 10.2307/1912557.

36. Cai M, Xie Y, Topol EJ, Al-Aly Z. Three-year outcomes of post-acute sequelae of COVID-19. Nat Med. 2024;30(6):1564–73. Epub 20240530. doi: 10.1038/s41591-024-02987-8. PubMed PMID: 38816608; PMCID: PMC11186764.

37. Sherif ZA, Gomez CR, Connors TJ, Henrich TJ, Reeves WB. Pathogenic mechanisms of post-acute sequelae of SARS-CoV-2 infection (PASC). Elife. 2023;12. Epub 20230322. doi: 10.7554/eLife.86002. PubMed PMID: 36947108; PMCID: PMC10032659.

38. Danielle RCS, Débora DM, Alessandra NLP, Alexia SSZ, Débora MCR, Elizabel NV, Felipe AM, Giulia MG, Henrique PR, Karen RMB, Layane SB, Leandro AB, Livia CM, Raquel SRT, Lorena SCA, Lyvia NRA, Mariana TR, Matheus CC, Vinícius DPV, Yasmin MG, Iúri DL. Correlating COVID-19 severity with biomarker profiles and patient prognosis. Sci Rep. 2024;14(1):22353. Epub 20240927. doi: 10.1038/s41598-024-71951-w. PubMed PMID: 39333538; PMCID: PMC11436624.

39. Diray-Arce J, Conti MG, Petrova B, Kanarek N, Angelidou A, Levy O. Integrative Metabolomics to Identify Molecular Signatures of Responses to Vaccines and Infections. Metabolites. 2020;10(12). Epub 20201130. doi: 10.3390/metabo10120492. PubMed PMID: 33266347; PMCID: PMC7760881.

40. Hu T, Liu CH, Lei M, Zeng Q, Li L, Tang H, Zhang N. Metabolic regulation of the immune system in health and diseases: mechanisms and interventions. Signal Transduct Target Ther. 2024;9(1):268. Epub 20241009. doi: 10.1038/s41392-024-01954-6. PubMed PMID: 39379377; PMCID: PMC11461632.

41. Raghubeer S, Matsha TE. Methylenetetrahydrofolate (MTHFR), the One-Carbon Cycle, and Cardiovascular Risks. Nutrients. 2021;13(12). Epub 20211220. doi: 10.3390/nu13124562. PubMed PMID: 34960114; PMCID: PMC8703276.

42. Moll S, Varga EA. Homocysteine and MTHFR Mutations. Circulation. 2015;132(1):e6–9. doi: 10.1161/circulationaha.114.013311. PubMed PMID: 26149435.

43. Ozonoff A, Schaenman J, Jayavelu ND, Milliren CE, Calfee CS, Cairns CB, Kraft M, Baden LR, Shaw AC, Krammer F, van Bakel H, Esserman DA, Liu S, Sesma AF, Simon V, Hafler DA, Montgomery RR, Kleinstein SH, Levy O, Bime C, Haddad EK, Erle DJ, Pulendran B, Nadeau KC, Davis MM, Hough CL, Messer WB, Higuita NIA, Metcalf JP, Atkinson MA, Brakenridge SC, Corry D, Kheradmand F, Ehrlich LIR, Melamed E, McComsey GA, Sekaly R, Diray-Arce J, Peters B, Augustine AD, Reed EF, Altman MC, Becker PM, Rouphael N. Phenotypes of disease severity in a cohort of hospitalized COVID-19 patients: Results from the IMPACC study. EBioMedicine. 2022;83:104208. Epub 20220808. doi: 10.1016/j.ebiom.2022.104208. PubMed PMID: 35952496; PMCID: PMC9359694.

