## supplemental material for "An Allele of the MTHFR one-carbon metabolism gene predicts severity of COVID-19"

Affiliations
1 Department of Pathology, Boston Children's Hospital, Boston, MA 02115, USA
2 Harvard Medical School, Boston, MA 02115, USA
3 Core Facilities, Medical University of Vienna, Austria
4 *Precision Vaccines Program*, Department of Pediatrics, Boston Children's Hospital, Boston, MA, 02115, USA
5 Research Computing, Department of Information Technology, Boston Children's Hospital, Boston, MA 02115, USA
6 Yale School of Medicine, New Haven, CT 06510, USA
7 Yale School of Public Health, New Haven, CT 06510, USA
8 Harvard Medical School, Boston, MA 02115, USA
9 Channing Institute of Medicine, Brigham and Women's Hospital, Boston, MA 02115, USA
10 Broad Institute of MIT & Harvard, 415 Main St, Cambridge, MA 02142, USA
### Corresponding authors (equal contribution)

#### Supplementary figures

**Figure S1. Integrated analysis of global and targeted plasma metabolite profiling of the IMPACC cohort.**

A A schematic outlining the scope of the global and targeted metabolomics assays as part of phases A and B of the IMPACC study[22]. Circles are depicted at scale to represent the number of samples analyzed in each assay and each phase. Filled color-coded area within the circles represents the number of samples analyzed by targeted metabolomics, and open area outlines represent the number of samples analyzed by global metabolomics. Phase A global n = 1055; Phase B global n = 2146; Phase A targeted n = 199 ; Phase B targeted n = 303

B Pearson correlation analysis on log-transformed and Pareto-scaled data for the indicated metabolites, comparing the same samples analyzed by both global and targeted metabolomics platforms. The Pearson correlation coefficient (r) and p-values are provided for each metabolite. global n = 2146; targeted n = 303

C Heatmap analysis of differential metabolites between batches 1 to 3 of targeted metabolomics with universal plasma sample (UPS) as associated quality control and mock samples. Analysis was performed on the MetaboAnalyst 6.0 platform, post log-transforming and Pareto scaling mean-centered data.

D Ordinal regression analysis from global metabolomics for methionine, cysteine, SAM and taurine metabolism pathway among all trajectory groups (TG 1,2,3,4,5). Significantly different metabolites ($p<0.01$, coefficient $>0$) are highlighted in magenta. n=1055

**Figure S2. Relative changes of metabolites in the methionine pathway are correlated with COVID-19 severity**

A Glycine levels from global and targeted metabolomics. Data were mean-centered, log-transformed, and Pareto-scaled. Left or right panels are depicting the same data organized either by trajectory groups (TG1-5) or by visit (V1,2,4 and 6) respectively. A two-way Anova (trajectory comparisons) or a mixed-effects (visit comparisons) analysis were performed, and corresponding p-values are indicated where *= $p<0.01$; **= $p<0.001$; ***= $p<0.001$; all remaining comparisons were not significant and were omitted for clarity. Global metabolomics n=2146; Targeted metabolomics n=199. TG – trajectory group. V- visit.

B As for A but depicting serine levels.

C S-adenosylhomocystein (SAH) levels from global metabolomics. Data are treated and represented as in A except SAH was detected only in the global metabolomics platform.

**Figure S3. Relative changes of metabolites in the of methionine pathway are correlated with COVID-19 severity**

A Schematic of one-carbon metabolism with focus on the enzyme MTHFR. Abbreviations are: DHF - dihydrofolate; THF -tetrahydrofolate; 5-meTHF – 5-methyl THF; 5,10-CH-THF – 5,10-methenyl THF; 5,10-$CH_2$-THF – 5,10-methylene THF; SAM – S-Adenosyl methionine; SAH – S-adenosylhomocysteine. Downstream one-carbon dependent reactions are indicated.

B Schematic of the hypomorphic *MTHFR* C677T and A1298C alleles (left panel) as well as the residual activity for the corresponding protein mutants (right panel). C677T allele, depicted in dark purple, is at the rs1801133 locus, and is the cause of the A to V amino acid substitution; A1298C allele, depicted in magenta, is at the rs1801131 locus and is the cause of the E to A amino acid substitution.

C Distribution of *MTHFR* C677T allele across the population as reported by Li et al and others[31, 32] in comparison to the IMPACC cohort. GG – wild type allele, GA – C677T heterozygous, AA – C677T homozygous and hypomorph allele.

D, E Distribution of *MTHFR* C677T allele across disease severity trajectory groups in the full IMPACC cohort (D), and in the targeted metabolomics cohort (E). TG – trajectory groups.

F Relative abundance by mass spectrometry of methionine, SAH, and 5-methyl THF. Methionine and SAH were measured using our global metabolomics platform, and 5-methyl THF using our targeted metabolomics. Data was mean-centered, log-transformed, and Pareto-scaled. Data is stratified by *MTHFR* allele and visit, after all trajectories were combined. A two-way Anova was performed, “ns” not significant. For methionine and SAH: V1, GG n = 480; V1, GA n = 345; V1, AA n = 129; V2, GG n = 304; V2, GA n = 204; V2, AA n = 82; V4, GG n = 164; V4, GA n = 116; V4, AA n = 36; V6, GG n = 153; V6, GA n = 102; V6, AA n = 31;
For 5-methyl THF: V1, GG n = 39; V1, GA n = 28; V1, AA n = 6; V2, GG n = 39; V2, GA n = 28; V2, AA n = 6; V4, GG n = 39; V4, GA n = 28; V4, AA n = 6; V6, GG n = 39; V6, GA n = 28; V6, AA n = 6;

**Figure S4.** ***MTHFR*** **allele frequency and effects across the IMPACC cohort**

A Frequency of trajectory groups (left panel) or mortality (right panel) across patients with the corresponding *MTHFR* alleles. rs1801133 refers to C677T, rs1801131 refers to A1298C.

B Hazard ratio analysis for *MTHFR* C677T (rs1801133) and A1298C (rs1801131) alleles. Univariate Cox proportional hazards model using the wildtype allele group as a reference. Survival was defined as the time from hospital admission to either death or time of the last follow-up. The p-values of the log-rank test were used to assess significance. For C677T: GG – wild type allele, GA – heterozygous allele, AA – homozygous hypomorph allele. For A1298C: TT – wild type allele, TG – heterozygous allele, GG – homozygous hypomorph allele

C Comparison of AIC and log-likelihood values for the Likelihood ratio test (LRT) results for model comparison in A. The inclusion of the three metabolites SAH, methionine and methionine-sulfoxide, improved model performance compared to the genetic-only (null) model as seen through lower AIC and log-likelihood values for the full model, while adding genetic information to the metabolite-based model did not enhance predictive power.

**Figure S5. Perturbation of methionine metabolism correlated with PASC status.**

A Levels of the indicated metabolites across *MTHFR* alleles comparing minimal and combined PASC clinical outcomes. Corresponding Wilcoxon rank sum p-values are indicated where *= $p<0.05$; all remaining comparisons were not significant and are not depicted.

**Figure S1**

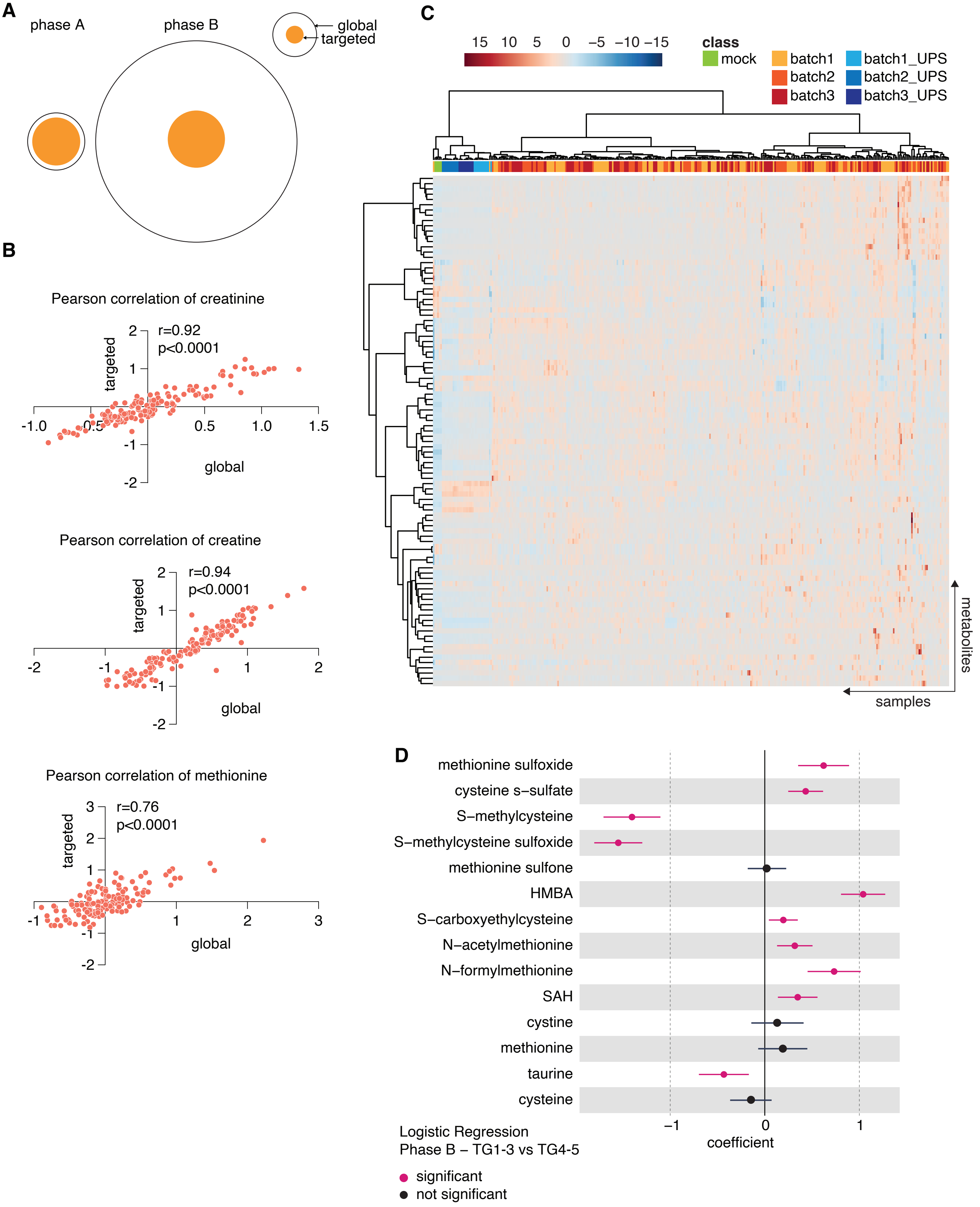
A
phase A
phase B
global
targeted
B
Pearson correlation of creatinine
r=0.92
p<0.0001
targeted
global
Pearson correlation of creatine
r=0.94
p<0.0001
targeted
global
Pearson correlation of methionine
r=0.76
p<0.0001
targeted
global
C
15 10 5 0 −5 −10 −15
class
mock
batch1
batch2
batch3
batch1_UPS
batch2_UPS
batch3_UPS
metabolites
samples
D
methionine sulfoxide
cysteine s−sulfate
S−methylcysteine
S−methylcysteine sulfoxide
methionine sulfone
HMBA
S−carboxyethylcysteine
N−acetylmethionine
N−formylmethionine
SAH
cystine
methionine
taurine
cysteine
coefficient
Logistic Regression
Phase B – TG1-3 vs TG4-5
significant
not significant

#### Figure S2

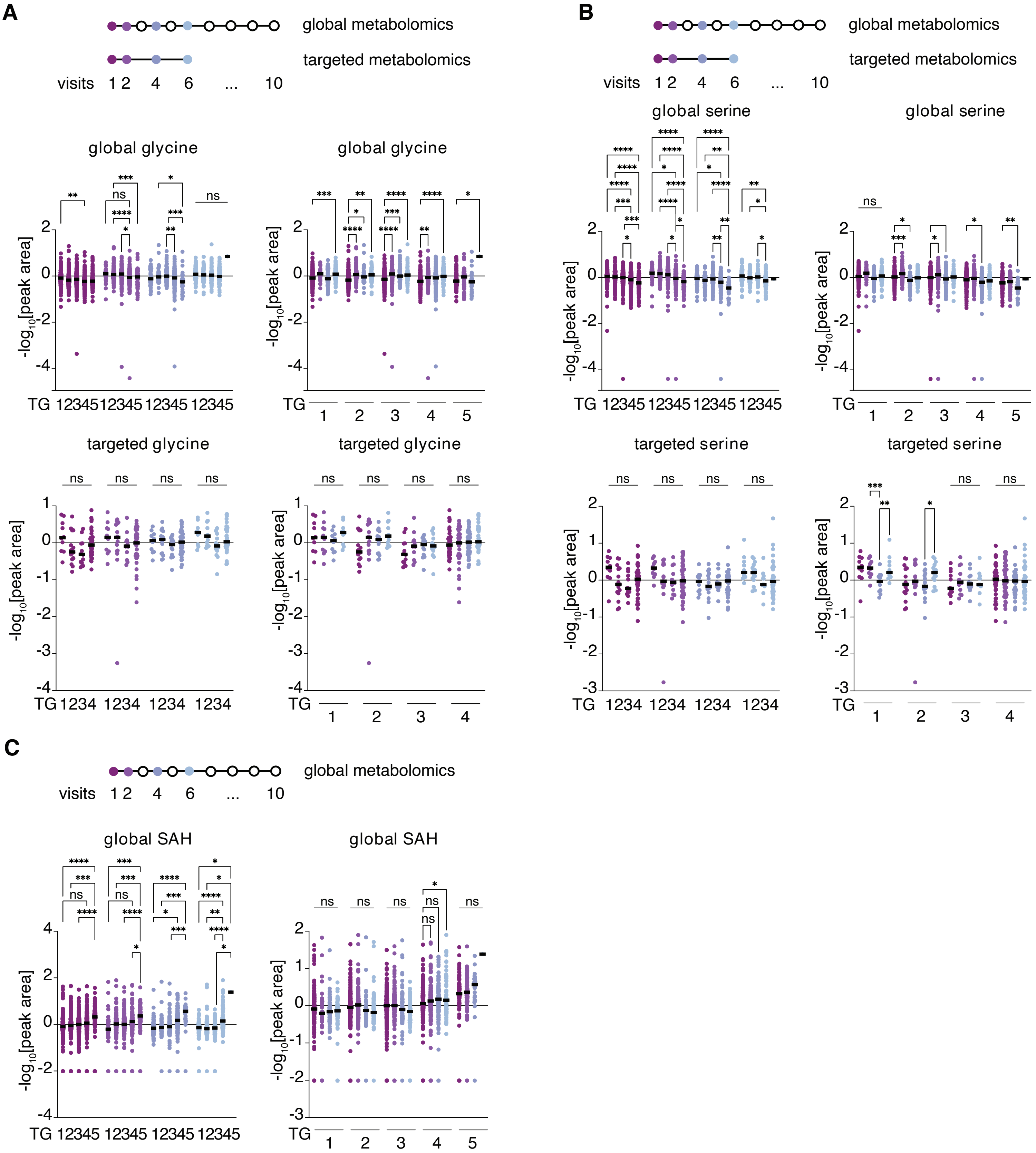
A
global metabolomics
targeted metabolomics
visits 1 2 4 6 ... 10
global glycine
global glycine
targeted glycine
targeted glycine
-log10[peak area]
TG 12345 12345 12345 12345
TG 1 2 3 4 5
TG 1234 1234 1234 1234
TG 1 2 3 4
B
global metabolomics
targeted metabolomics
visits 1 2 4 6 ... 10
global serine
global serine
targeted serine
targeted serine
-log10[peak area]
TG 12345 12345 12345 12345
TG 1 2 3 4 5
TG 1234 1234 1234 1234
TG 1 2 3 4
C
global metabolomics
visits 1 2 4 6 ... 10
global SAH
global SAH
-log10[peak area]
TG 12345 12345 12345 12345
TG 1 2 3 4 5

Figure S3

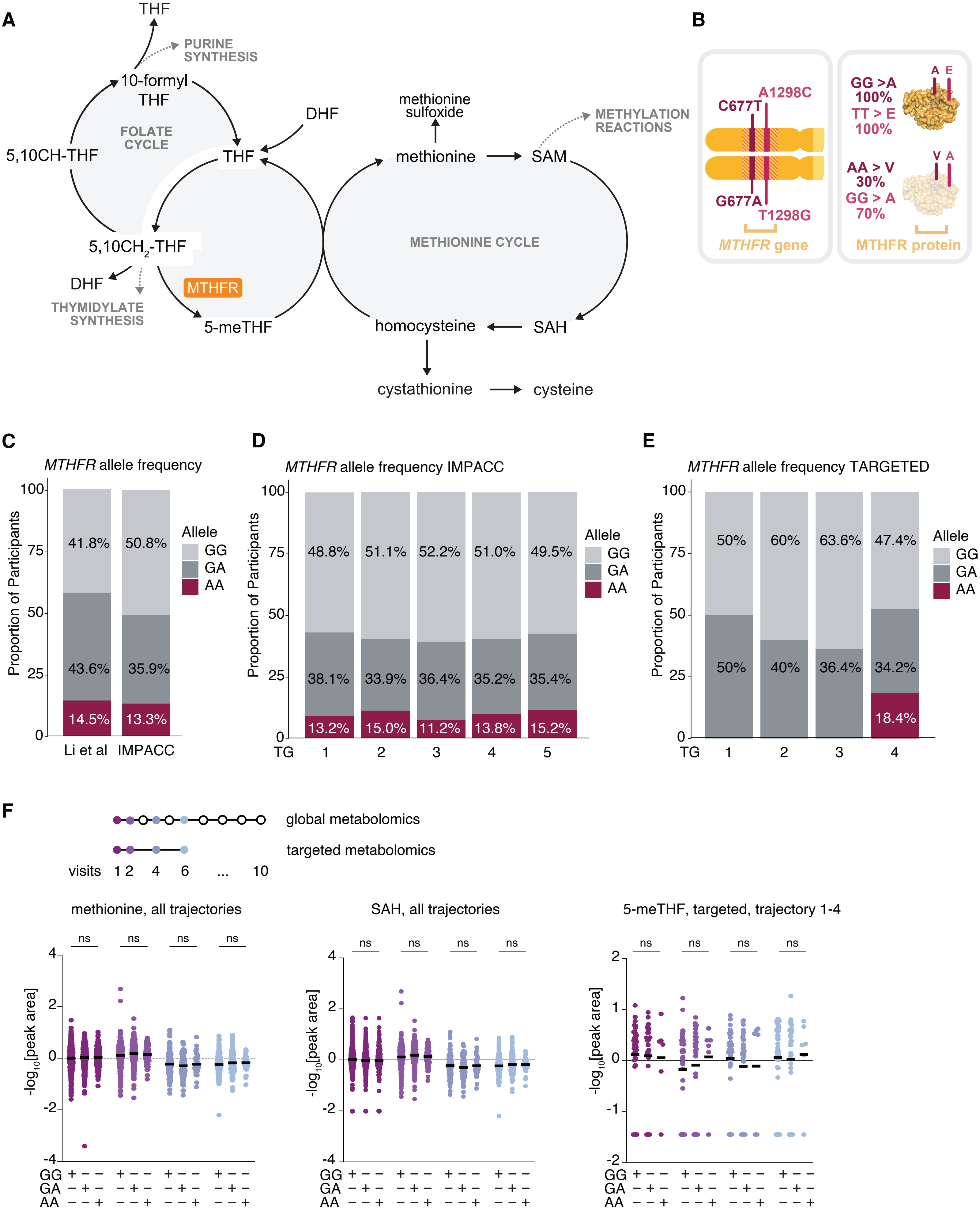

A
THF
PURINE SYNTHESIS
10-formyl THF
FOLATE CYCLE
5,10CH-THF
THF
DHF
5,10CH2-THF
DHF
THYMIDYLATE SYNTHESIS
MTHFR
5-meTHF
methionine sulfoxide
methionine
SAM
METHYLATION REACTIONS
METHIONINE CYCLE
homocysteine
SAH
cystathionine
cysteine
B
C677T
A1298C
G677A
T1298G
MTHFR gene
A E
GG >A 100%
TT > E 100%
V A
AA > V 30%
GG > A 70%
MTHFR protein
C
MTHFR allele frequency
Proportion of Participants
Allele
GG
GA
AA
41.8% 50.8%
43.6% 35.9%
14.5% 13.3%
Li et al IMPACC
D
MTHFR allele frequency IMPACC
48.8% 51.1% 52.2% 51.0% 49.5%
38.1% 33.9% 36.4% 35.2% 35.4%
13.2% 15.0% 11.2% 13.8% 15.2%
TG 1 2 3 4 5
E
MTHFR allele frequency TARGETED
50% 60% 63.6% 47.4%
50% 40% 36.4% 34.2%
18.4%
TG 1 2 3 4
F
global metabolomics
targeted metabolomics
visits 1 2 4 6 ... 10
methionine, all trajectories
SAH, all trajectories
5-meTHF, targeted, trajectory 1-4
ns
-log10[peak area]
GG
GA
AA

**Figure S4**

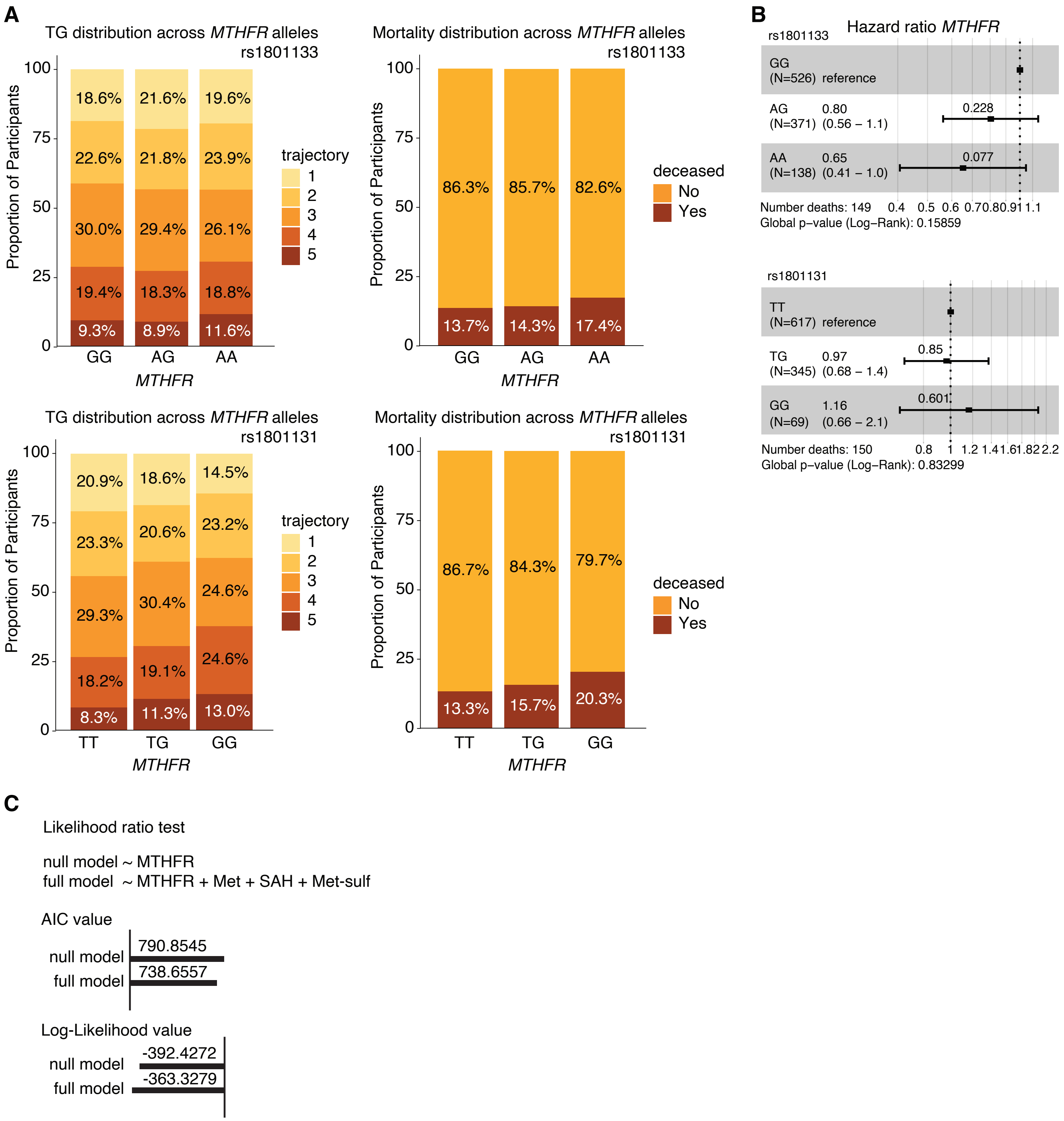
A
TG distribution across MTHFR alleles rs1801133
Proportion of Participants
100
75
50
25
0
18.6%
21.6%
19.6%
22.6%
21.8%
23.9%
30.0%
29.4%
26.1%
19.4%
18.3%
18.8%
9.3%
8.9%
11.6%
GG
AG
AA
MTHFR
trajectory
1
2
3
4
5
Mortality distribution across MTHFR alleles rs1801133
86.3%
85.7%
82.6%
13.7%
14.3%
17.4%
deceased
No
Yes
TG distribution across MTHFR alleles rs1801131
20.9%
18.6%
14.5%
23.3%
20.6%
23.2%
29.3%
30.4%
24.6%
18.2%
19.1%
24.6%
8.3%
11.3%
13.0%
TT
TG
GG
Mortality distribution across MTHFR alleles rs1801131
86.7%
84.3%
79.7%
13.3%
15.7%
20.3%
B
Hazard ratio MTHFR
rs1801133
GG (N=526) reference
AG (N=371) 0.80 (0.56 – 1.1) 0.228
AA (N=138) 0.65 (0.41 – 1.0) 0.077
Number deaths: 149
0.4 0.5 0.6 0.7 0.8 0.9 1 1.1
Global p–value (Log–Rank): 0.15859
rs1801131
TT (N=617) reference
TG (N=345) 0.97 (0.68 – 1.4) 0.85
GG (N=69) 1.16 (0.66 – 2.1) 0.601
Number deaths: 150
0.8 1 1.2 1.4 1.6 1.8 2 2.2
Global p–value (Log–Rank): 0.83299
C
Likelihood ratio test
null model ~ MTHFR
full model ~ MTHFR + Met + SAH + Met-sulf
AIC value
null model
790.8545
full model
738.6557
Log-Likelihood value
null model
-392.4272
full model
-363.3279

#### Figure S5

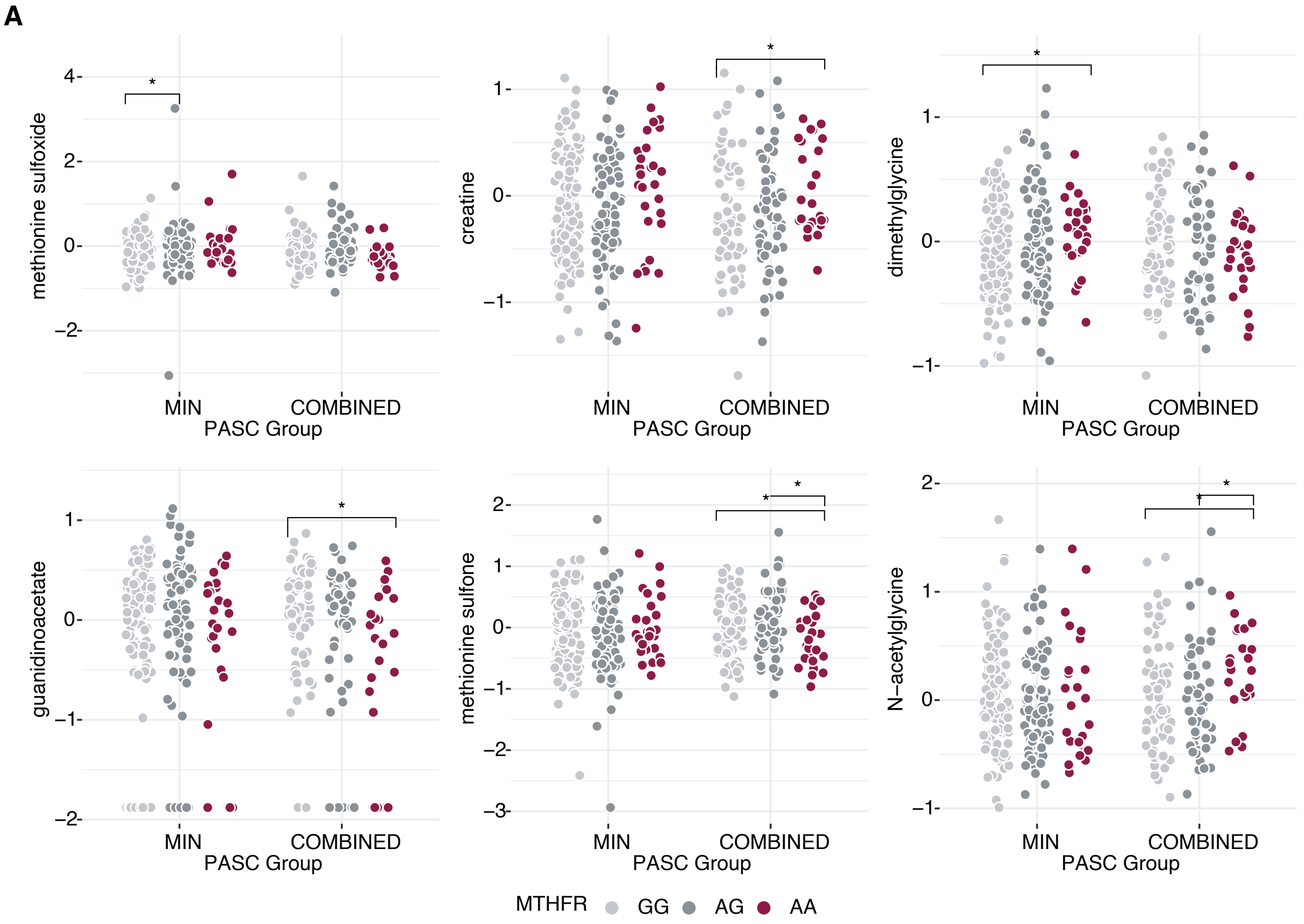
A
methionine sulfoxide
creatine
dimethylglycine
guanidinoacetate
methionine sulfone
N−acetylglycine
MIN
COMBINED
PASC Group
MTHFR
GG
AG
AA
